# Strict Physical Distancing May Be More Efficient: A mathematical argument for making the lockdowns count

**DOI:** 10.1101/2020.05.19.20107045

**Authors:** Scott Sheffield, Anna York, Nicole Swartwood, Alyssa Bilinski, Anne Williamson, Meagan Fitzpatrick

## Abstract

COVID-19 created a global public health and economic emergency. Policymakers acted quickly and decisively to contain the spread of disease through physical distancing measures. However, these measures also impact physical, mental and economic well-being, creating difficult trade-offs. Here we use a simple mathematical model to explore the balance between public health measures and their associated social and economic costs. Across a range of cost-functions and model structures, commitment to intermittent and strict social distancing measures leads to better overall outcomes than temporally consistent implementation of moderate physical distancing measures. With regard to the trade-offs that policymakers may soon face, our results emphasize that economic and health outcomes do not exist in full competition. Compared to consistent moderation, intermittently strict policies can better mitigate the impact of the pandemic on both of these priorities for a range of plausible utility functions.

## 1 Introduction

COVID-19 is a respiratory disease caused by the novel coronavirus SARS-CoV-2. Since identification in December 2019, SARS-CoV-2 has spread rapidly around the world, with substantial morbidity and mortality: as of May 7, 2020, there were more than 3,847,000 confirmed cases and 270,000 confirmed deaths globally [1]. To slow the spread of disease and prevent overwhelming health care services, many governments initiated non-pharmaceutical interventions (NPIs) that called for a high degree of physical distancing. While effective in dampening disease spread, these measures have had dramatic economic and social effects. The impact on unemployment has been particularly marked; in the week ending May 7, 2020, the US Department of Labor reported 33 million initial unemployment claims over the past 7 weeks [2]. Due to these costs, there have been increasing calls to reduce physical distancing measures, leaving policymakers with difficult trade-offs.

Previous research has argued that one-time interventions will be insufficient to maintain control of the COVID-19 pandemic and highlighted the need for long-term application of physical distancing measures [3, 4, 5]. At face value, it may seem reasonable to assume that adopting “moderate measures” may both slow the spread of disease *and* simultaneously permit some level of normality which may help mitigate against some of the adverse effects listed above. Nevertheless, it remains an open question whether it is optimal to apply strong, intermittent measures or long-term moderate measures. One previous paper advocated for a “severe lockdown” which tapers gradually based on an optimal control solution to an SIR model [6]. Another proposed tapering lockdown based on age [7]. However, both papers used an SIR model, which does not consider the incubation period of the disease, and considered only a single utility function. By contrast, several epidemiological papers have noted that intermittent lockdowns may offer a route to prevent critical care capacity from being overwhelmed while allowing for periods of greater economic activity [4, 8]. Other optimal control papers have proposed both suppression and maintenance strategies [5] or intermittent lockdowns [9]. However, these papers modeled only disease control and did not explicitly consider non-disease costs.

In this paper, we combine a simple epidemiological model with a model of costs associated with lockdown to compare intermittent and moderate lockdown strategies. We explore mathematically what various assumptions about the utility function would imply about the optimal form of the long-term strategy. We show that for a range of utility functions, committing to coordinated but intermittent stricter physical distancing measures leads to better outcomes than consistent implementation of moderate physical distancing measures over the same time period. While we use the example of “lockdowns,” these findings could apply to a range of NPIs, including school closures and business re-openings. Our objective is not to recommend a specific optimal strategy for a particular setting. Rather, our work highlights a general finding: that under a range of plausible assumptions, stricter measures are more efficient relative to their cost. This understanding is of particular importance for public acceptance of these measures, as well as helping to inform and support the best policy decisions during this uncertain time.

## 2 Methods

### 2.1 Exponential growth model

We developed a simplified deterministic model that simulates a pandemic with exponential growth within a closed population. Our model assumes (à la Reed-Frost [10]) that if an individual contracts the virus during week *n*, then the individual will be exposed but not infectious for the remainder of week *n*, infectious throughout the duration of week *n* + 1, and no longer infectious (or safely quarantined) at all times after week *n* + 1.

*R*_0_ represents the basic reproductive number, which is the average number of secondary infections arising from a single infectious case in a completely susceptible population. Although in general the relationship between *R*_0_ and the weekly growth rate depends on the length of the incubation and infectious periods [11], the very simple assumptions above imply that

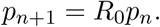

where *p_n_* represents the fraction of people actively infectious during week *n*. With physical distancing measures in place, our new equation becomes:

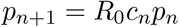

where *c_n_* represents the fraction of normal social exposure on average during the *n*th week, due to social restrictions, and *R*_EFF_(*n*) = *R*_0_*c_n_* is the *effective reproductive number* in the *n*th week.

For clarity of exposition, this analysis includes two simplifications of common epidemiological models:

1. **S remains constant** Our model investigates long-term management of low-level disease. Therefore, we assume that the total infection rate (over the period studied) is small compared to the susceptible population. Similar assumptions are commonly used to approximate early-stage SEIS or SEIR, when nearly everyone is susceptible and one obtains a roughly linear ODE involving only the infected states *E* and *I* [11]. This approach (keeping *S/N* near 1 and *R/N* near zero) is used in a related COVID-19 analysis evaluating intermittent strategies through a dynamic transmission model [8].
2. **Time is discrete** Our simplified model assumes that, if an individual contracts the virus during week *n*, then the individual will be exposed but not infectious for the remainder of week *n*, infectious throughout the duration of week *n* + 1, and either safely quarantined or no longer infectious at all times after week *n* + 1. This weekly “lag time” mimics the function of the exposed class *E* within transmission models. Although more rigid than a model solved in continuous time, it is similar to discretized dynamic models that require policies to be fixed one week at a time, as detailed further in Appendices F and G.

We connect this epidemiological model to economic outcomes through a utility function *U*(*c*) which encodes the cost of reducing the weekly disease transmission rate by a factor of *c*. The function *U* can in principle be determined experimentally, as more countries try different lockdown approaches and try to observe both how expensive and how effective they are. An advantage of our simplified approach is that an empirical study of the function *U* can be conducted without making additional assumptions about the disease.

### 2.2 Model parameters

#### 2.2.1 *R*_0_

We assume that with no government restrictions (but with general public awareness and voluntary behavior changes), each infectious individual would infect 2.5 additional people. As a weekly growth factor, this value is within the estimated range of *R*_0_ for COVID-19 in many countries [12, 13, 14]. It is also within the range of the *R*_0_ values that arise from fitting SEIR models to empirical data, though we stress again that *R*_0_ values do not correspond exactly to weekly growth factors in SEIR [11]. Our timescale, which corresponds to roughly 3.5 days prior to infectiousness and a week of infectivity also roughly corresponds to COVID-19 [15].

#### 2.2.2 Physical distancing

Due in part to the social exposure of essential workers, *c_n_* = 0 will never be achieved. We assume that *c*_min_ = 0.16 is the lowest possible value of *c_n_*. This reduction aligns with the observation that the extreme measures taken in Wuhan reduced the effective reproductive number to 0.32 [16]. The latter number would correspond to *p_n_*_+1_/*p_n_* being slightly smaller than 0.4, also similar to New Zealand’s estimates after successful mitigation [17]. We assume that reductions beyond this point are not practical.

Therefore, in the intermittent model the strictest possible measures (*c_n_* = *c*_min_ = .4/ *R*_0_ = .16) result in *p_n_*_+1_ = .4 *p_n_*, and the mildest reasonable measures (*c_n_* = 1) result in *p_n_*_+1_ = 2.5 *p_n_*. In the “consistently moderate” lockdown, intermediate measures (*c_n_* = 1/ *R*_0_ = .4) would result in *p_n_*_+1_ = *p_n_*. While these values are within the range of current COVID-19 estimates [18, 19, 20], they represent simplified rounded estimates and are not proposed to be representative of any particular location. We consider variations of these in sensitivity analysis to consider less strict “down” periods and milder “up” periods. We also discuss how, in some cases, measures like contact tracing might reduce *P* without substantially affecting *U*, thus allowing greater gains at lower costs.

### 2.3 Utility function

We consider a range of forms for the utility function *U*, with a particular focus on *U*(*α*) = *c^α^* for *α* ϵ (0,1]. The simple case *U*(*c*) = *c* assumes that overall utility is reduced by the same proportion as social contacts. Other choices of *α* account for the fact not all contacts are equal: some are more *costly* to eliminate than others and some are more *important* to eliminate than others (e.g., those most likely to involve both an infected and a susceptible individual) and hence one would not expect the slope of *U* (which encodes the *marginal* cost of reducing the percentage of new infections) to be constant. The value *α* encodes the rate at which this marginal cost increases (if *α* < 1) or decreases (if *α* > 1) as more contacts are eliminated.

In order to estimate the appropriate functional form for the costs of lockdown, existing work (reviewed in Appendix A) typically has taken one of two approaches: top-down or bottom-up. In the first approach, one estimates overall GDP contraction after lockdown measures were imposed; in the latter, one sums up the estimated costs to individuals from each activity forgone. For our base case, we adopt the “bottom-up approach.” In a simplified example, suppose our primary policy were to close some number of factories. If *k* open factories produce *k* times as much utility (and also *k* times as many contacts) as one open factory, then both utility and disease transmission rates would be linear functions of the number remaining open, so that could we write *U*(*c*) = c (“linear policy tool”). On the other hand, if a random fraction of the population were required to wear burdensome but perfectly effective masks, then the utility *u* could be the fraction of people *not* required to wear them. If non-mask-wearer interactions were proportional to *u*^2^, we could then write *c* = *u*^2^ so that 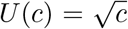 (“quadratic policy tool”). Vaccination and lockdowns are sometimes modeled similarly [21, 22, 23]. If during a lockdown only a *u* fraction of workers are considered *essential* and allowed to work, then one could argue either that *c* = *u* (if the essential workers have just as many contacts per worker as usual) or that *c* = *u*^2^ (if they have only *u* times as many contacts per worker as usual), depending on the nature of the lockdown. In a complex society combining many different kinds of policies, *U*(*c*) = *c^α^* for some *α between* 1/2 and 1 might therefore be reasonable.

For technical readers, our analysis is further guided by the following observations:

1. **Restricting the utility function domain:** Some activities have very low value relative to the amount of contact they require (e.g., because they have substitutes, such as online meetings, that don’t require contact). Some interventions (hand-washing, masks) may also be low cost (compared to school or business closings). It is safe to assume that during an “up period” these effective-but-inexpensive measures (“low-hanging fruit”) would remain in place, but we consider these costs as outside of the scope of our utility function. In other words, we assume that these inexpensive measures are already taken in the *c* = 1 scenario.
2. **Diminishing returns:** It is possible that once the extremes are eliminated, there remains in the interval [*c*_min_, 1] a broad range of social activity that is roughly equal in value, so that *U* is approximately linear on this range. However, it is reasonable to guess that even within the interval [*c*_min_, 1], some contacts are less costly to cut than others, so that there are diminishing returns to cost. This would suggest *U* is *concave*, like *U*(*c*) = *c^α^* with *α* < 1.
3. **Crowding effects:** For another perspective, one may imagine that the number/type of economic activities is fixed, and that the only question is how to distribute them temporally. If *α* = 1 then the utility of “full activity one week, zero activity next week” equals the utility of “half activity both weeks.” In practice though, maintaining safety standards might be harder or more expensive during a full activity week (e.g., if activities have to be moved to late or early hours to avoid crowding). Choosing *α* < 1 would account for the associated cost.
4. **Population effects:** We do not explicitly address population inhomogeneity in this paper. However, we note that if only a *u* fraction of workers are allowed to work, and after a short period of time most of the infections are among *these* workers, and if these workers have nearly as much contact as usual (as they work hard to provide for those at home) then we might actually find c close to 1 even when *u* is small. In particular, we might find *c* > *u* (as when *α* > 1).
5. **Accelerating returns:** While the first contacts one eliminates are less likely to be costly, they are also more likely to be “redundant.” In other words, if a person inhales infected droplets multiple times during the course of week, and each of these exposures is sufficient to transmit the disease, then removing only one exposure does not prevent infection; one needs further measures before the impact is felt. Because of this phenomenon, some disease transmission models actually suggest a convex *U*. An example involving SEIR on a low-degree social network appears in Appendix G.2. For a simpler example, imagine Activity A and Activity B would each expose 10 people if allowed to proceed, but that there is some overlap; say 3 people would be exposed in both places. Then cancelling Activity A alone only prevents 7 infections while cancelling both activities prevents 17 infections. If both activities are equally valuable, then the second cancellation achieves more relative to its cost than the first. This phenomenon might play a significant role if new infections occur largely among close associates of individuals with high viral loads, and if “redundant exposures” among this vulnerable group are common; see Appendix G. This is another reason to consider larger *α* values.

Based on the above, we consider three ranges of a values:

1. **Convex:** *U*(*c*) = *c^α^* for *α* > 1.
2. **Moderate:** *U*(*c*) = *c^α^* for *α* ϵ (0,1].
3. **Ultra-concave:** *U*(*c*) = - *c^α^* for *α* < 0.

We focus primarily on the moderate scenario. That is, our baseline assumption is that diminishing returns play a larger role than accelerating returns (so that *α* ≤ 1) but not so large that they lead to *α* < 0. We stress that *U* depends *both* on the variation in economic value attached to different activities *and* on the model governing the disease transmission; Appendix G considers a range of *U* obtained by varying both parameters.

These results can also be generalized to utility functions outside of the *U*(*c*) discussed above. As discussed in Appendix F, for *any* twice-differentiable function *U* we can define

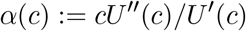

and say that *U* is convex, moderate or ultra-concave on an interval based on the value of *α*(*c*) on that interval. The basic results in this paper (about the optimality of intermittent strategies) apply to intervals on which *U* is convex or moderate. (Ultra-concavity is equivalent to concavity on a logarithmic scale, as Appendix F explains.)

### 2.4 Strategy comparison

We assume that a vaccine will be available in a known period of time (72 weeks). Therefore, our focus is on successfully managing the 72 week “holding period.” Specifically, we aim to minimize the total infection rate (*P*) by implementing control measures while simultaneously maximizing productive activities or utility (*U*), which in turn minimizes the social and economic “cost” associated with these control measures. Total infection rate is given as

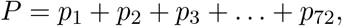

and total utility is

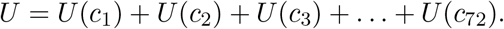

We presume that policymakers want to keep cases below a threshold. For example, hospital capacity (e.g., the number of regular staffed beds or ICU beds) is a key limitation in successful disease control. This is important both in terms of direct ability to treat COVID-19 patients and more generally as a proxy for whether the health system is overburdened and unable to adequately treat patients with other conditions. We therefore assume, based on American Hospital Association surveys, that if the infection rate exceeds one case per 1000 (*H* = 0.001), then the hospitals will be above capacity [24]. For simplicity, we will use “hospital capacity” as an absolute cut-off and aim to keep the infection rate below this threshold. (This constraint prevents *P* from becoming large enough to produce substantial herd immunity.) We also assume that once the virus reaches about 4 cases per million, it cannot be reduced any further, regardless of any additional physical distancing measures (e.g., because there will always be a few infections from unrecognized lingering illness, or from outside the country). We seek to choose *c_n_* in order to maximize *U* subject to the constraints: *p_n_* ϵ [.000004, .001] for all *n* and *c_n_* ϵ [*c*_min_, 1] for all *n*. For illustration, we compare the following two strategies:

1. **Steady “less extreme” physical distancing:** Adopt moderate distancing (*c_n_* = .4) every week for the whole 72 weeks. Note that in this case *p_n_*_+1_ = 2.5 *c_n_ p_n_* = *p_n_* (i.e. the number of people affected remains constant from one week to the next; the effective *R*_0_ is 1), and hospital capacity remains steady at *H*.
2. **Intermittent strict lockdown:** Start off with an extreme total lockdown (*c_n_* = .16) for six weeks while the virus percentage decreases to its lowest level. Then relax the measures and allow normal life to continue (*c_n_* = 1) until new patients reach the hospital capacity threshold. Then lockdown again and repeat this cycle until the 72 weeks are up.

We also note underlying mathematical properties that make intermittent distancing preferable to steady moderation.

### 2.5 Extension to SEIR model

We also present results from a standard SEIR (or SEIS) epidemiological framework with susceptible (S), exposed (E), infectious (I), and recovered (R) compartments. We focus on the early-phase linearized ODE (i.e., the large *S*, small *R* limit) in which only the exposed and infectious populations vary in time [11]. We simulate the same tradeoffs under a range of parameters, to reflect different values for the effective reproduction number during the up periods and the down periods.

In each case the mean incubation period is 4 days and the mean infectious (but not isolated) period is set to 4 days, with durations that are Erlang with shape parameter 2, as in [8]. Erlang distributions have been applied to COVID-19 in many places [25, 26, 27] and adopting an Erlang distribution with parameter *k* is equivalent to subdividing a compartment into *k* sub-compartments (which leads in our case to four states total: *E*_1_,*E*_2_,*I*_1_,*I*_2_, similar to [8]). One of the early papers on the incubation time (based on cases in China) fit the incubation time to an Erlang distribution with shape parameter 6 [28], which is equivalent to dividing *E* into six sub-states *E*_1_, *E*_2_,…, *E*_6_. However, this was a model of time to symptom appearance, which is different from (and easier to directly measure than) time to infectiousness.

The relative lengths of the cycles were adjusted to produce peaks of approximately *H* = 0.001. In this setting we focus on the ratio *R*_EFF_ / *R*_0_ during the *n*th period. The steady moderation strategy corresponds to fixing *R*_EFF_ = 1 in each period. We also briefly discuss mathematical properties that determine the optimal solution in this case, contrasting it to simple exponential model. We defined *U* by integrating (*R*_EFF_ / *R*_0_)*^α^* instead of *c^α^*. (We discuss a way to make sense of *U* as a function of c in this setting in Appendix G.)

## 3 Results

For intuition, we first consider the model’s behavior under the two most extreme scenarios: complete inaction and complete lockdown. Complete inaction (*c_n_* = 1 for all *n*) would result in the infection growing by a factor of 2.5 weekly until most people are infected. It yields *U* = 72, the maximum value, indicating that utility is “as normal” but generates an unacceptably large *P* and rapidly exceeds hospital capacity. And it is worth noting that, in the real world, complete inaction would not yield the maximal *U*, because a large number of infections and associated deaths would cause societal and economic disruption as a byproduct. At the other extreme, a completely strict lockdown (*c_n_* = .4 for all *n*) minimizes *P* but yields a low *U*, indicating unacceptably large societal and economical costs.

Figure 1 compares two competing strategies in the exponential model: a “consistently moderate” lockdown corresponding to *R*_EFF_ = 1, and an alternation between six-week strict lockdown periods and six-week periods without major restrictions. While we do not focus here on optimizing prevalence beyond H, the hospitalization threshold, we note that the first measure yields *P* = .072 (so 7.2 percent of the population is ultimately infected) while the second yields *P* ≈ .014 (1.4 percent). If we adopt the simple utility function *U*(*c*) = c (which assumes that the cost of eliminating some amount of social exposure is proportional to the amount) then we find *U* = .4 · 72 = 28.8 under steady moderation and *U* = .16 · 36 + 36 = 41.76 under intermittent lockdown. If instead we take *U*(*α*) = *c^α^* for some *α* > 0 then steady moderation yields *U* = 72(.4)*^α^* while intermittent lockdowns yield *U* = 36(.16)*^α^* + 36. In fact, for any utility function of this form, strict intermittent distancing would dominate steady, moderate distancing because of a mathematical theorem called the alpha-geometric mean inequality.^1^

**Figure 1:**
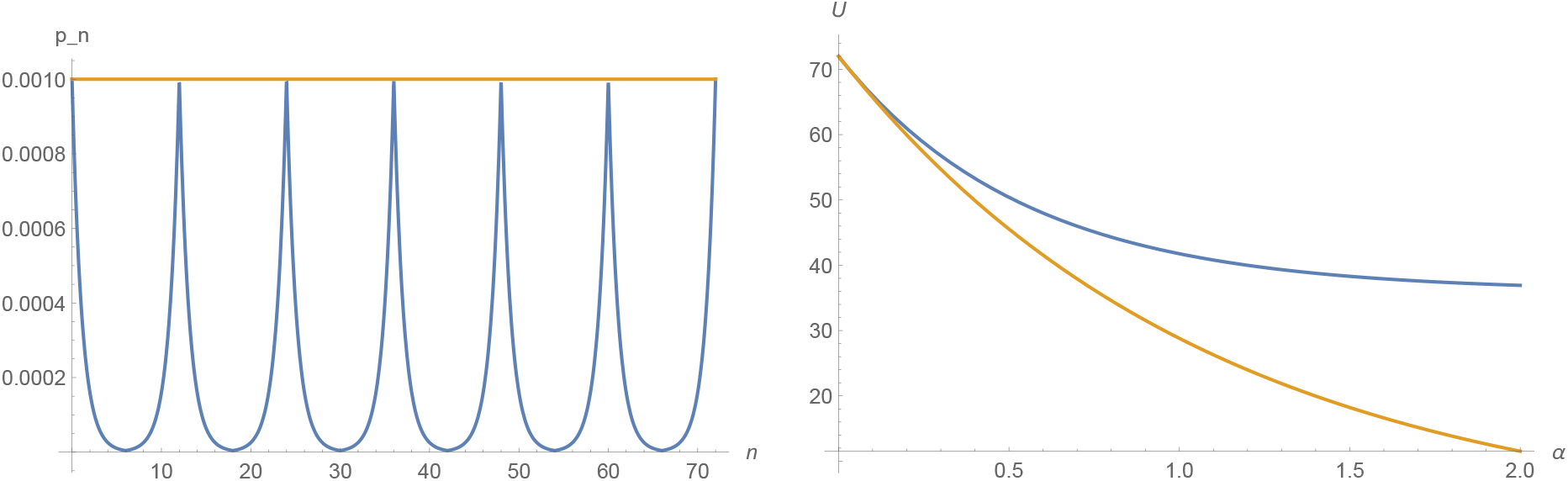
Left: comparison of *p_n_* with steady moderation (orange) and intermittent lockdown measures (blue). Right: utility under steady moderation (orange) and intermittent lockdown (blue) when *U*(*c*) = *c^α^*.

With a simple exponential model, an optimal solution would involve short cycles. For example, another strategy is even better than the one illustrated in Figure 1: namely, first 6 weeks down, then 30 1-week-up-1-week-down cycles, and then 6 weeks up. This would achieve the same *U* as the strategy in Figure 1 but with a lower *P*. However, we will explain in see Figure 3 that there is in fact a cost (not accounted for in the model above) to increasing the number of cycles, which would make it less efficient to have many very short cycles.

When we expand this analysis to SEIR models, Figure 2 illustrates that (at least for *α* > 1/2) intermittent strict lockdowns always resulted in larger *U* and smaller *P* than steady moderation. However, gains decrease substantially when either physical distancing is less strict during down periods or productivity during up periods is smaller. In Appendix G.2, we discuss mathematically why intermittent strategies dominate steady moderation in these models, noting that the utility gains may be diminished if the up and down periods are very short, as shown in Figure 3.

**Figure 2:**
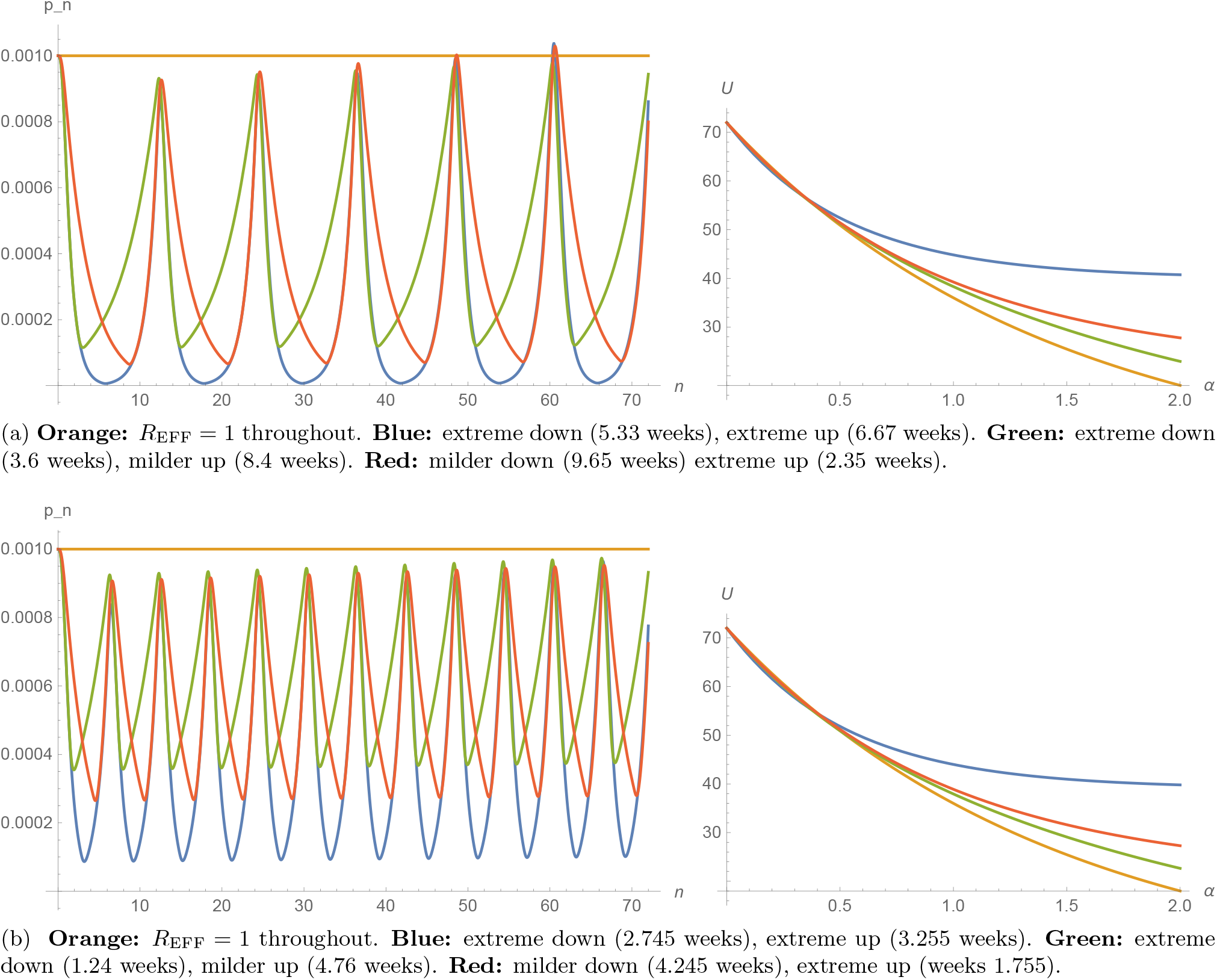
Left figures show evolution under various intermittent strategies with 12-week periods (above) or 6-week periods (below). In each case either *R*_EFF_ = 2 (extreme up), *R*_EFF_ = 1.25 (milder up), *R*_EFF_ = 1 (moderate), *R*_EFF_ = .7 (milder down) or *R*_EFF_ = .3 (extreme down). Right figures show corresponding *U* as function of *α*, with same colors. Mean incubation and infection times set to 4 days in a linearized ODE derived from two-state Erlang SEIR. At time zero, *E*_1_, *E*_2_, *I*_1_, and *I*_2_ are all equal; the plots illustrate *I*_2_.

**Figure 3:**
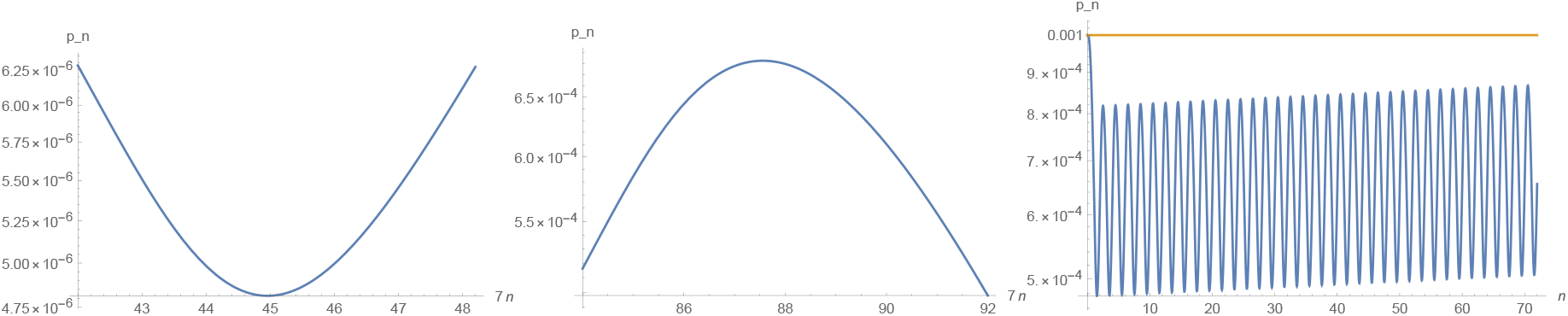
Zooming in on the upturns (left) and downturns (center) of an analog of Figure 2 in which *R*_EFF_ = 2.2 during up and .33 during down periods (which leads to roughly steady peaks if one has exactly 6-week up and 6-week down periods). The transitions (viewed on a log scale) are “rounded,” unlike the sharp transitions in (the log scale version of) Figure 1. After removing a lockdown (left) it takes about **6 days** for to return to the original height and the steady state increasing slope. After imposing a lockdown (center) it takes nearly **8 days** for to return to the original height and the steady state decreasing slope. Alternating 6.15 days up, 7.85 days down (right) keeps the peaks roughly steady. But it requires more lockdown days overall than the longer-period alternation (42 days up, 42 days down) in Figure 2b. Nonetheless, even this rapid oscillation still yields a larger *U* than steady moderation if *U*(*c*) = *c^α^* and *α* > .63.

## 4 Discussion

In this paper we explored the utility of different physical distancing lockdown cycles. We use a simplified transmission model to evaluate the effect of different lengths of strict measures (lockdowns) and “free” periods on both disease progression and non-disease utility. Our analysis shows that (unless *U* is ultraconcave) the consistently moderate lockdown is worse, for both public health and utility, when compared to intermittent cycles of strict physical distancing followed by periods of (relative) normality. As others have noted, if regular lockdowns are simple to implement and predictable for people to follow, they may be a useful tool, particularly if it were difficult to maintain steady moderation near the threshold, necessitating lockdowns to ensure containment even when attempting steady moderation [9].

These results provide a utility-based justification of epidemiological papers that recommend intermittent lockdowns [4, 29, 3] and characterize the conditions under which intermittent lockdowns are likely to be preferable to steady moderation. However, our results differ from optimal control models [23, 7] that recommend steady moderation. Some of this difference is explained by their use of an SIR model without an incubation period, which allows for a type of “continuous compounding” not seen in our models, effectively leading to an ultra-concave U; see Appendix G.

### 4.1 Contextualizing our results in the rapidly evolving pandemic

In the fast-moving COVID-19 pandemic, it can be difficult to contexualize model findings in light of developments that may arise, for example, in testing and contact tracing programs, therapeutics, and new understanding about seroprevalence. We show how our results may be impacted by these in Figure 4, which illustrates that combating the virus can be viewed metaphorically as walking the wrong way on a moving sidewalk. Inaction results in a steady drift to the left (toward high death rates and greater immunity), while distancing measures involve walking or running to the right at different speeds. As one drifts toward the left endpoint, the fraction of infected individuals grows large enough so that the susceptible population can no longer be treated as constant, and some level of herd immunity is acquired. Near the right endpoint, contact tracing and targeted quarantine may prove less costly. For example, suppose that when an individual tests positive *all* of the 1000 or so *remotely* connected individuals are immediately quarantined. As extreme as that would be, if testing were widespread and the number of weekly confirmed positives were low (say 4 per million) it would still be less disruptive than a national lockdown.

**Figure 4:**
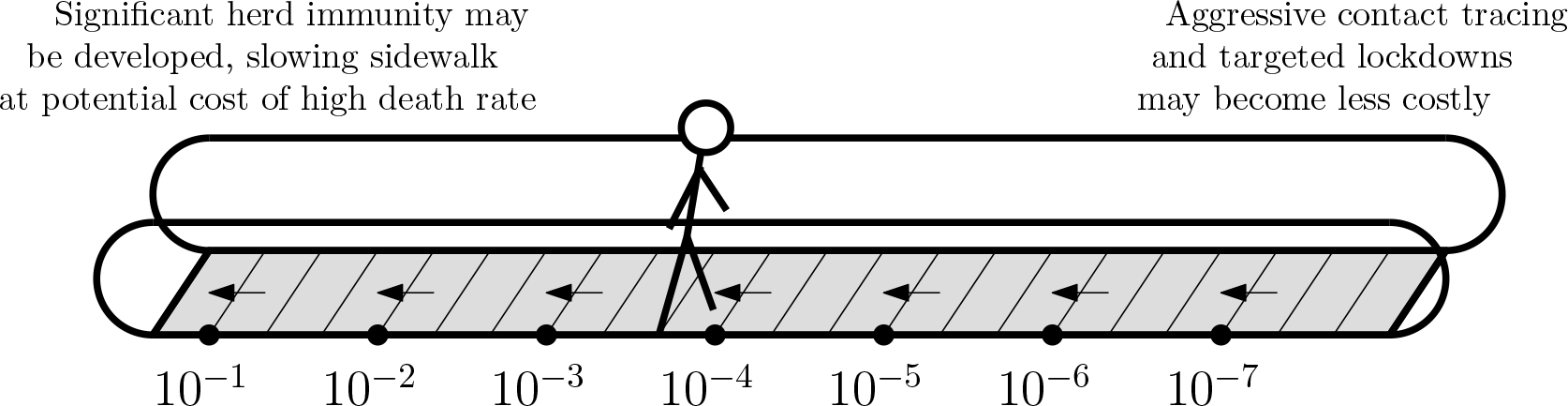
Numbers indicate disease prevalence. Inaction yields exponential growth (steady drift left).

The model in this paper does not address either endpoint and focuses only on the most efficient way to navigate the middle range, finding that (for a range of qualitatively plausible utility functions) alternating speeds is more efficient than maintaining position in a steady way. If it turns out that maintaining position near the right end is less costly than in the middle, then the subsequent peaks in Figures 1 and 2 might be unnecessary, and a larger value of *U* might be possible. Similarly, if it proves possible to acquire significant herd immunity while shielding the vulnerable (e.g., with prophylactics or treatment) then this would lead to slower leftward drift, which could be navigated in a less costly way. Within the context of SEIR, if 20 percent of the population were immune, then an infectious person would only have 80 percent as many contacts with non-immune people, which would suggest an *R*_EFF_ only 80 percent as large as otherwise.

We stress that in a society that acquires significant herd immunity (deliberately or otherwise), it would still be possible to adopt an intermittent strategy later on, and at that point it would be possible to do so with shorter lockdowns and longer up periods. The model in this paper applies *after* one has determined not to allow a large number of additional infections but *before* it has become possible to inexpensively maintain position on the right.

Finally, despite evidence for temperature and climate variation [30, 31, 32], it is unclear if the pathogen’s transmissibility will exhibit seasonal effects. In contrast to [4], we do not assume seasonal changes in the disease spread; that is, our model will produce the same magnitude of effect no matter where one places the lockdowns in the calendar year. If the SARS-CoV-2 virus displays the same seasonality as SARS or the influenza virus, this would “slow the sidewalk” during the summer months, and the required lockdown durations would vary seasonally.

### 4.2 Caveats and limitations

Our work is also subject to a number of limitations.

1. **Strict distancing is very strict:** The “moderate” measures required to achieve *R*_0_ ≈ 1 might still be tremendously strict on an objective scale. In many countries, one cannot say for sure whether it will even be possible to pursue a strategy like the one proposed here before new tools are assembled, and the benefits of intermittent strategies are much lower if *R*_DOWN_ cannot be reduced considerably below 1. Likewise, if individuals take additional precautions (beyond policy recommendations) or are hesitant to engage in economic activity during the “up” periods then the benefits may be diminished as well. It might be easier to initiate productive up periods when the infection rates are legitimately low (so that people know they have less to fear). It may also pose additional logistical challenges to implement an intermittent strategy, due to both transition costs not modeled here and to risks associated with underestimating transmission during “up” periods.
2. **Inhomogeneity:** Limited COVID-19-specific information about the distribution of incubation times or infectiousness patterns has led us to consider basic Erlang SEIR approximations. If it turns out that a significant subgroup of people remain infectious for very long periods of time, this would make it harder to reduce infection rates quickly. Likewise, we do not account for subgroup differentiation in infectious rates or other random fluctuations or superspreader events.
3. **Utility function uncertainty:** Due to absent empirical data, it is hard to assess the impact of “accelerating return” and “diminishing return” effects. In particular, we cannot rule out the possibility that *U* is ultra-concave, which would lead to steady moderation being an optimal strategy; see Section F for more details. Moreover, we have not treated the possibility that *U* itself may change gradually over time, due e.g. to the increased availability of personal protective equipment; the improvement of test-and-trace technology; or the gradual improvement of safety protocols within schools and businesses.

## 5 Conclusions

In light of all of the caveats above, we stress that we are not arguing that the specific pattern that appears in Figure 1 is likely to appear in our future. Nonetheless, this simple model illustrates a few key points:

1. **Full measures beat half measures:** In certain situations, our normally healthy aversion to extreme measures can lead us badly astray. Reducing the virus 99 percent one month achieves the same reduction as *two* 90-percent-reduction months, or *nearly seven* 50-percent-reduction months. *If* expensive measures are being undertaken, then going the extra mile to make those measures *as airtight as possible* might significantly increase their effectiveness, and thereby greatly decrease the amount of time they have to be in place. For a considerable range of utility functions, it appears to be better to alternate between stricter periods and more relaxed periods than to try to produce a single sustainable policy.
2. **Coordinating lockdowns may increase returns:** It is often better to combine similarly costly restrictions in the same time period than to space them out over different periods. Instead of only asking “Which kinds of work are inessential or doable from home?” policy makers should also ask “Which kinds of work can be *staggered* (e.g., 60 hour weeks during up periods, no work during down periods, assuming that this does not substantially change spacing and other safety requirements in place)?” The more that work can be staggered (packed into up periods, left out of down periods) the closer the down periods get to the *zero transmission* ideal, and the more impactful they become.
3. **Containment is ideal:** Although alternating between slower and faster speeds may be more efficient, swimming against an “exponential current” for 72 weeks is costly. An aggressive test-and-trace program (if successful) might be the only way to simultaneously achieve a low *P* and a high *U* and reduce the losses associated with physical distancing measures.

The simple models presented here support stricter, intermittent lockdowns compared to moderate, consistent distancing strategies. This evidence suggests that, for a certain range of utility functions, strict intermittent measures are more efficient for public health as well as social and economic well-being. To achieve optimal utility, the timing of distancing measures and the identification of permitted activities in “up” and “down” periods should be considered in future research. Successful implementation of intermittent lockdowns will not only require coordination and cooperation from the public, but additional, clear policy leadership and government financial support will be essential for the necessary adjustments to be made. As companies and individuals adjust to this new intermittent way of life, the cost of lockdowns and the corresponding utility will likely change. With new data every day, we urge for models to be updated and policy measures reviewed during this maintenance period. As stated previously, this work does not attempt to forecast or recommend any specific policy. Instead, we emphasize the potential for intermittent strategies to truly make lockdowns count.

## Data Availability

All data is generated from easily reproducible simulations, the source code (which expands on the Mathematica code presented and annotated in Appendix H) is provided as a Supplementary file.

## Appendix A A Brief Review of Literature on Lockdown Cost

As mentioned earlier, there are two ways to estimate the costs associated to a lockdown: “top down” (estimating overall GDP or economic sector contraction after lockdown measures are imposed) or “bottom up” (summing estimated costs to individuals from each activity forgone, for instance lost wages). Several papers follow the former approach, with [33] estimating that a flu pandemic which costs 1.4 million lives would reduce total output by almost 1 percent. They also showed that as the scale of the pandemic increases, so does the economic cost. Focusing on disruption to supply chains, consumption distortion and the financial market, [34] models the COVID-19 pandemic and estimates a GDP loss for the USA ranging from $16 billion if 1 percent of the population are infected and 0.02 percent die, to $1,769 billion if 30 percent are infected and 0.9 percent die. Others use data from past epidemics to calculate costs; during the 2003 SARS epidemic, the total loss to China’s economy due to reduced travel and tourism was estimated to be $25.3 billion [35]. The “bottom up” approach allows specific focus on physical distancing measures. A case study on the 2003 SARS outbreak in Toronto quantified quarantine costs as a function of administration costs, forgone daily wages of quarantined workers, and the number of contacts each infected person has, with estimated savings of $232 million Canadian dollars when compared to no containment mechanism [36].

## Appendix B Another normalization of the utility function

If one defines utility by *U*(*c*) = *c^α^* then it is automatically the case that the utility differences between different strategies involving *c* ϵ [*c*_min_, 1] become small as *α* → 0. In order to keep the difference from becoming trivial as *α* → 0 we can replace *U* by an affine function of *U* chosen so that 0 corresponds to the utility *U_min_* achieved when *c* = *c*_min_ throughout and 1 corresponds to the utility *U*_max_ achieved when *c* = 1 throughout. This is implemented in Figure 5 below, which better illustrates the extent to which consistent moderation is *better* when *α* is very small or negative. We stress that if the difference between the extreme values *U*_max_ and *U_min_* is very large (the equivalent of many trillions of dollars and/or millions of lives) then even differences that appear small (as a percentage of this difference) may be tremendously important.

**Figure 5:**
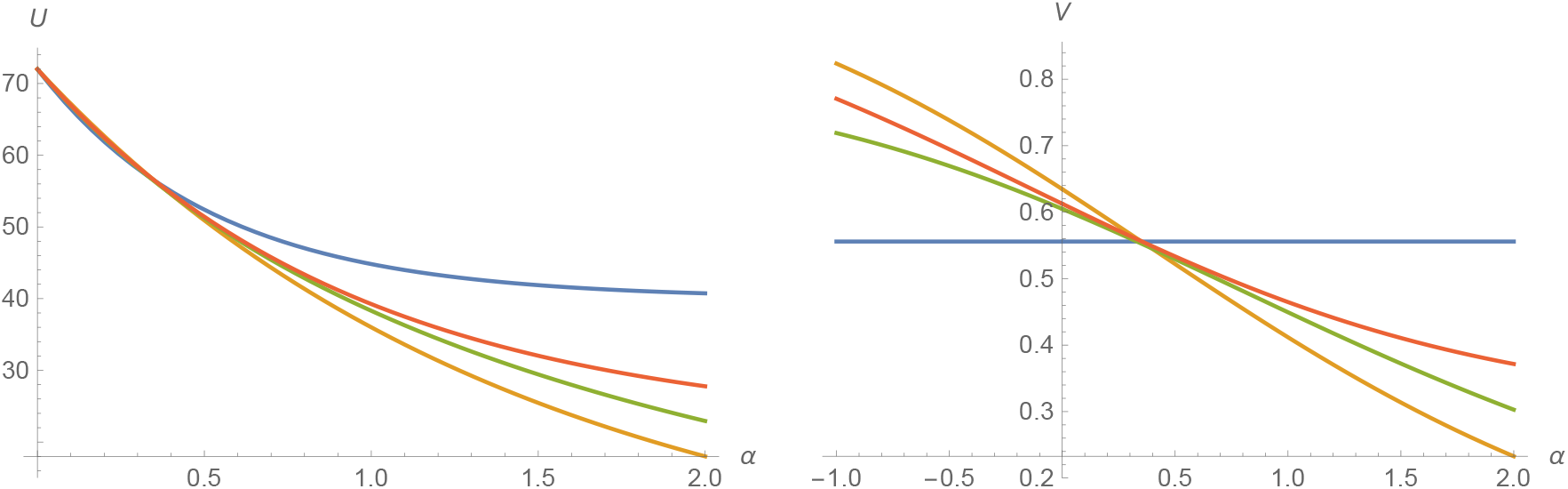
Left: utility chart from Figure 2. Right: same but with *U* replaced by 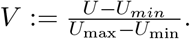.

## Appendix C Understanding the price of adding an extra cycle

Figure 6 uses the same parameters as Figure 3 and illustrates the full state vector (*E*_1_*, E*_2_*, I*_1_, *I*_2_) over the course of two 6-week-down-6-week-up cycles. The standard early-phase linearization of SEIR (or SEIS) with Erlang parameter 2 (see [8, 11]) is given by:

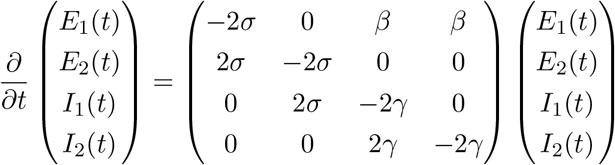

where *σ*^-1^ and *γ*^-1^ are the mean durations of the incubation and infectious periods and *R*_0_ = *β*/*γ* so that *β* = *γ R*_0_. If *σ* and *γ* are fixed, we then we write *m_R_* to denote the above matrix with *P* chosen so that *β*/*γ* = *R*. Here *R* represents the effective value *R*_EFF_.

**Figure 6:**
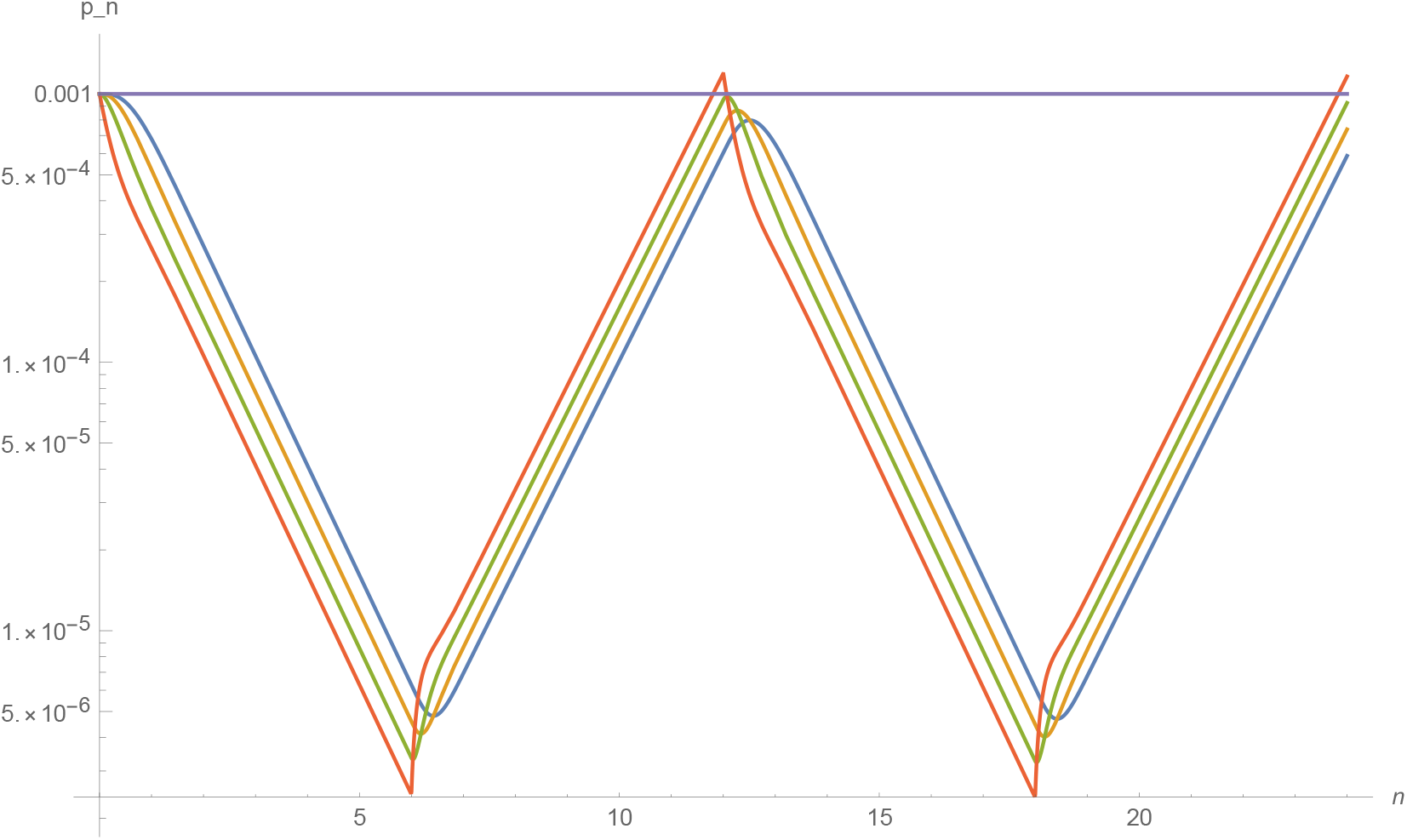
Same parameters as Figure 3: *R*_EFF_ = 2.2 during 6-week-long up periods and *R*_EFF_ = .33 during 6-week-long down periods, and periods of six weeks up, six week down, incubation/infection periods Erlang with mean of four days, shape parameter two. Two cycles (24 weeks) shown. All four states illustrated: *E*_1_(*t*) (red), *E*_2_ (*t*) (green), *I\*(*t*) (orange), and *I*_2_ (*t*) (blue).

When a policy change is made, so that *R*_EFF_ changes, the red curve (corresponding to *E*_1_) is the first to change direction: this is the “leading indicator” that the other curves lag behind. The fact that the space between the curves is roughly constant on the log scale (except for shortly after a policy change) corresponds to the fact that the ratios (*E*_2_/ *E*_1_ and *I*_1_/*E*_1_ and *I*_2_/*E*_1_) are roughly constant, which in turn corresponds to the fact that (*E*_1_, *E*_2_, *I*_1_, *I*_2_) (interpreted as a column vector) is close to a multiple of the Perron-Frobenius eigenvector of 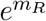, which is also the eigenvector of *m_R_* corresponding to the maximal real eigenvalue *λ_R_* [11].

The quantity *λ_R R_* is called the *Malthusian parameter* of *R*, and indicates the asymptotic slope of the lines in Figure 6 during a period when *R*_EFF_ = *R* [11]. We stress that unlike the basic reproduction number *R*_0_ (which describes the early-phase discrete exponential growth rate w.r.t. *generation number*) the Malthusian parameter encodes a continuum exponential growth rate w.r.t. *time*.

When the eigenvectors of *m_R_* are denoted by *v_j_*, we let *Q_R_* denote the projection operator that takes *v* = Σ*a_j_v_j_* to *a*_1_*v*_1_, where *v*_1_ is the eigenvector with dominant eigenvalue *λ_R_*. Note that

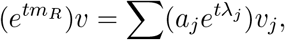

and that the RHS terms corresponding to *j* ≠ 1 grow exponentially more slowly in t than the leading term, which implies that

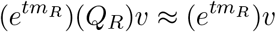

when *t* is large. Consider what happens between the 12 week mark and the 24 week mark of Figure 6. At the beginning, *v*_12_ is approximately an eigenvector of 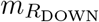 with *R*_DOWN_ = .33. But then (setting *t* = 6) we find

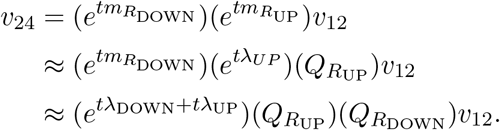

Let *C*(*R*_1_,*R*_2_) denote the log of the non-zero eigenvalue of 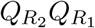 (and observe that *v*_12_ is approximately an eigenvector of this matrix), so that:

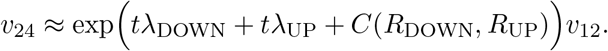

If the vector were simply growing at an exponential rate of *λ*_DOWN_ during down periods and *λ*_UP_ during up periods, then the above would hold without the C(*R*_DOWN_,*R*_UP_) term. The term *C*(*R*_DOWN_,*R*_UP_) represents an additional adjustment or “cost” associated to the switching of policies back and forth. Precisely, assuming all cycle lengths are reasonably long, 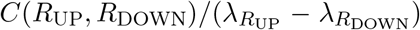approximates the amount of time one would have to swap from “up” to “down” in order to compensate for increasing the number of up-down cycles by one.

Let us work out this calculation in the simple example above. Using weeks as our unit, we have *σ* = *γ* = 7/4 and *β* = *R*_0_/*γ* so that, if .33 and 2.2 are the two effective *R*_0_ values, the corresponding two matrices become

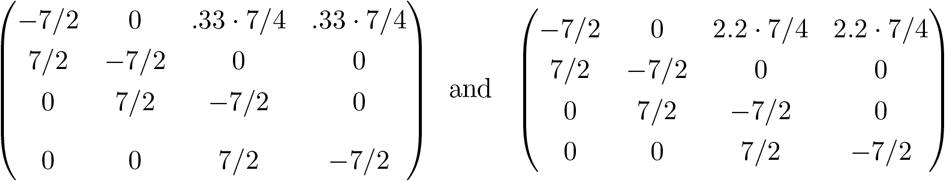

Entering these into an eigenvalue calculator we find 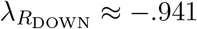 and 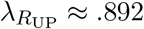.

The four ending values on the LHS side of Figure 7 exceed their counterparts on the RHS by about log 1.3068 ≈ .26758. Dividing by 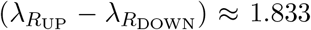 we obtain .146 which is reasonably close to 1/7. This suggests that adding an extra up-down cycle has about the same cost (in terms of its impact on the final values) as switching one day from down to up. (This cost per extra cycle would be different if the periods were short enough that could not be well approximated by an eigenvector at the end of each.) In Figure 3, switching from 6 cycles (of 12 weeks each) to 36 cycles (of 30 weeks each) means that one pays a price of about 30 extra days of lockdown (or 5/6 of a day per each two-week period) which is roughly what Figure 3 shows.

**Figure 7:**
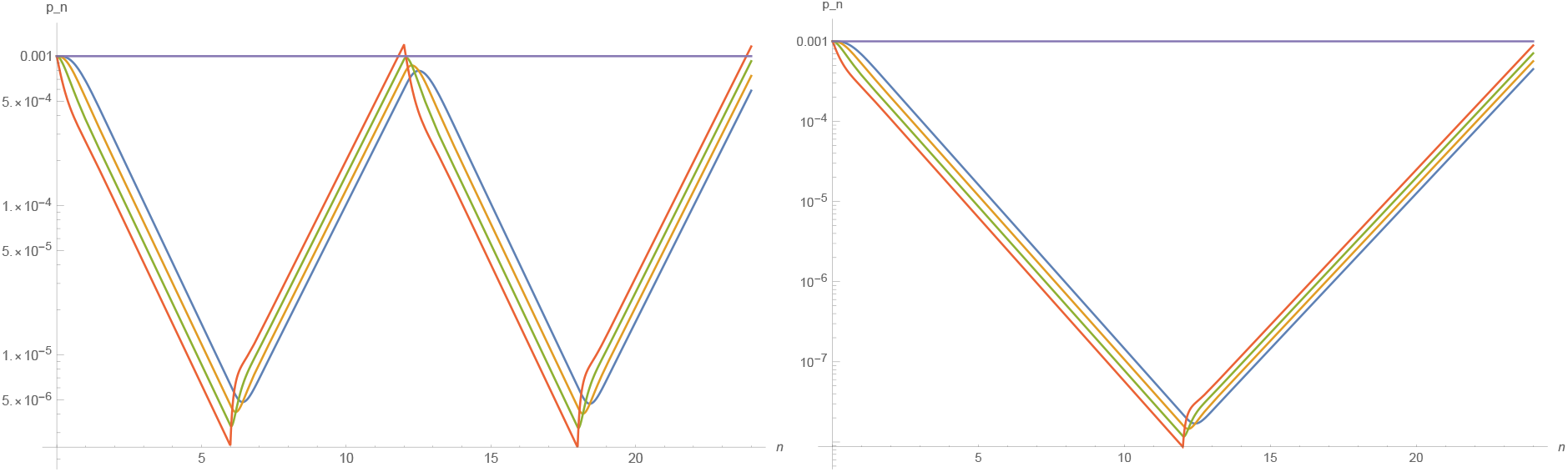
Same parameters as Figure 3: *R*_EFF_ = 2.2 during 6-week-long up periods and *R*_EFF_ = .33 during 6-week-long down periods, and periods of six weeks up, six week down, incubation/infection periods Erlang with mean of four days, shape parameter two. All four states illustrated: *E*_1_(*t*) (red), *E*_2_ (*t*) (green), *I*_1_(*t*) (orange), and *I*_2_(*t*) (blue).

In this example, the takeaway is that if the public prefers shorter cycle lengths (and hence a larger number of cycles), then this can be accommodated, but at the cost of about one extra day of lockdown for each extra cycle added.

## Appendix D Changing the Erlang parameter

An Erlang distribution with an integer *shape parameter k* and a positive real *rate parameter λ* (or equivalently a *scale parameter μ* = 1/ *λ*) is a probability density function defined for *t* ϵ [0, ∞) by

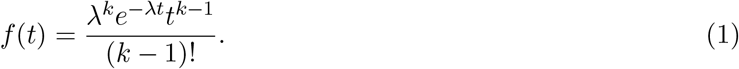

This is equivalent to a Gamma distribution, except that it comes with the extra requirement that *k* be an integer. When *k* = 1, (1) is the density function of an exponential random variable with rate *λ* (and expectation *μ* = 1/*λ*). For general *k*, (1) is the density function of a sum of *k* independent exponential random variables, each with rate *λ* (and expectation *μ* = 1/ *λ*); the overall sum then has expectation *kμ*.

The incubation period for COVID-19 was studied in [28], based on early data from China, and was fit to (among other things) an Erlang distribution with scale parameter *μ* = .880 and shape parameter *k* = 6 (which would correspond to a mean incubation time of 5.28) as shown in Figure 8. Per this distribution, the incubation period would have a low probability of being less than 2 days (about 2.85 percent) or greater than 14 days (about .15 percent).

**Figure 8:**
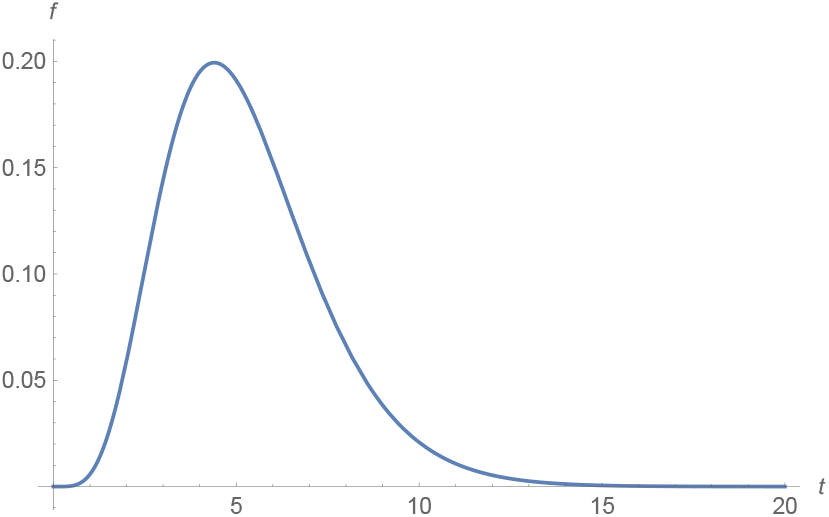
Erlang distibution with shape parameter *k* = 6 and scale parameter *μ* = .88.

However, the study in [28] considered only the time from first exposure until the development of *symptoms* among individuals who ultimately became symptomatic. We are interested in a harder-to-measure quantity: the time until a person becomes infectious (and subsequently the time between infection and isolation). In Figure 2, we implemented SEIR with incubation and infectious periods given by Erlang distributions with shape parameter of 2. However, in light of [28], one might propose a higher shape parameter (say *k* = 4 or *k* = 6) as potentially more realistic. Although it might be less realistic, one could also consider a shape parameter of 1 to correspond to the classical formulation of SEIR. The corresponding density functions would be as in Figure 9.

**Figure 9:**
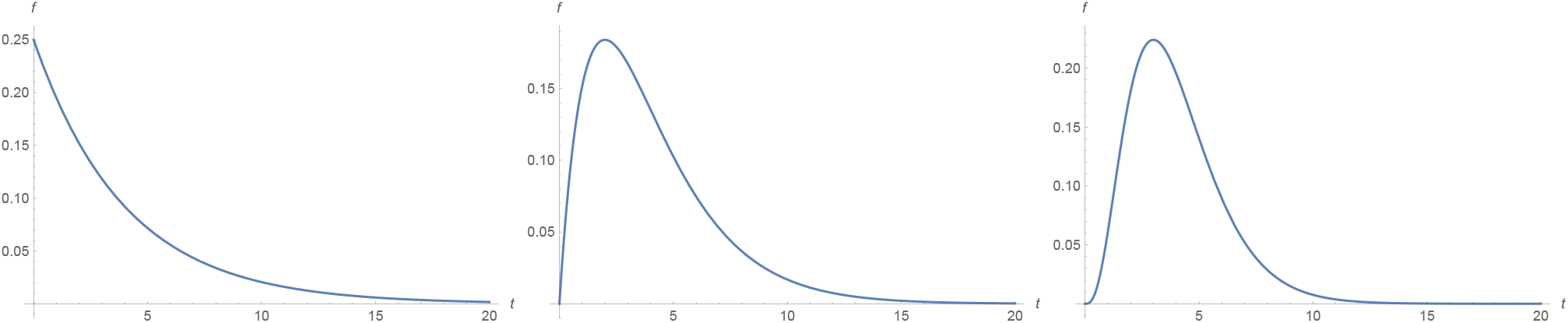
Erlang with mean *kμ* = 4 and shape parameter *k* = 1 (left), *k* = 2 (center) and *k* = 4 (right). The one on the right is more similar to incubation distributions observed empirically.

In a conventional SEIR or SEIS model, adopting an Erlang distribution for the law of the time an individual spends in state *E* is equivalent to replacing *E* with *k* separate states *E*_1_*, E*_2_*,…,E_k_* that one moves through sequentially. (Same for state *I*.) As Figure 10 illustrates modifying the Erlang parameter in the more realistic direction (to *k* = 4) strengthens the case for intermittent strategies (they become superior for smaller *α*); and Figure 11 illustrates that modifying it in the less realistic direction (to *k* = 1) weakens case for intermittent strategies (one requires a larger *α* for intermittent strategies to be superior).

**Figure 10:**
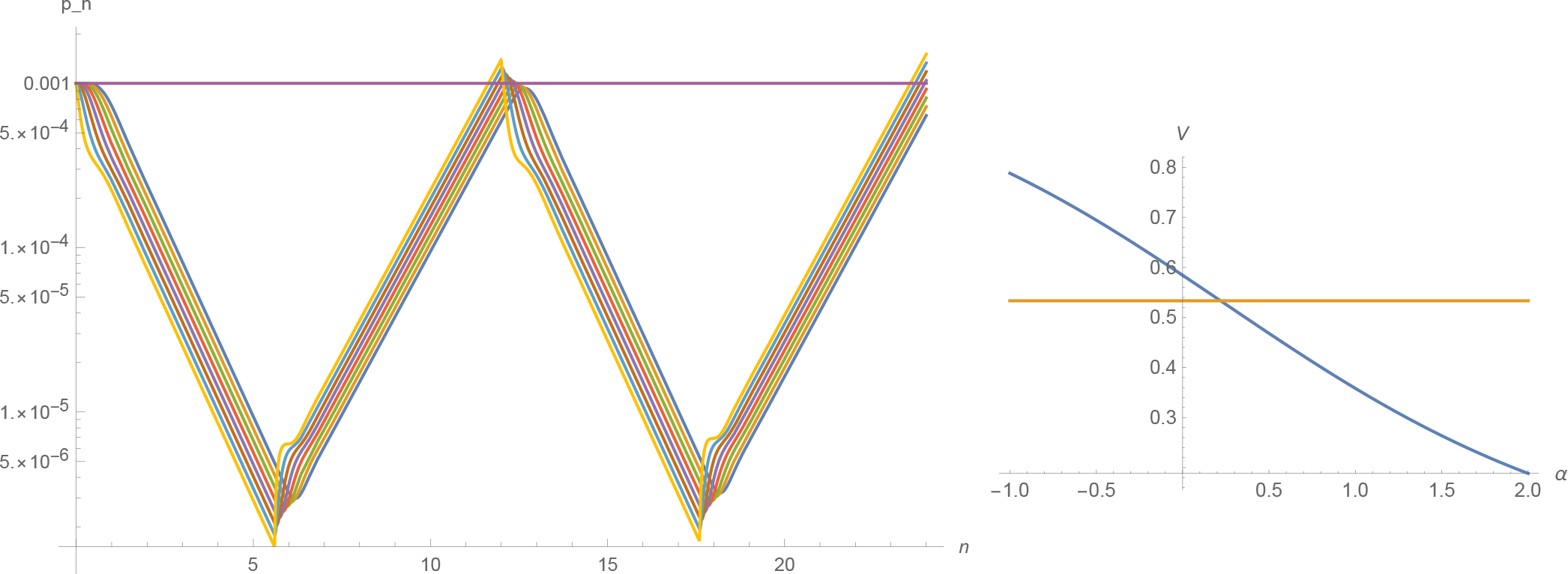
Left: same as Figure 6 except with Erlang shape parameter 4 instead of 2, and with cycles of 6.4 weeks up and 5.6 weeks down. All eight states illustrated: *E*_1_ (yellow), *E*_2_ (light blue), *E_3_* (brown), E_4_ (purple), *I*_1_ (red), *I*_2_ (green), *I_3_* (orange), and I_4_ (blue). Right: utility normalized as in Figure 5.

**Figure 11:**
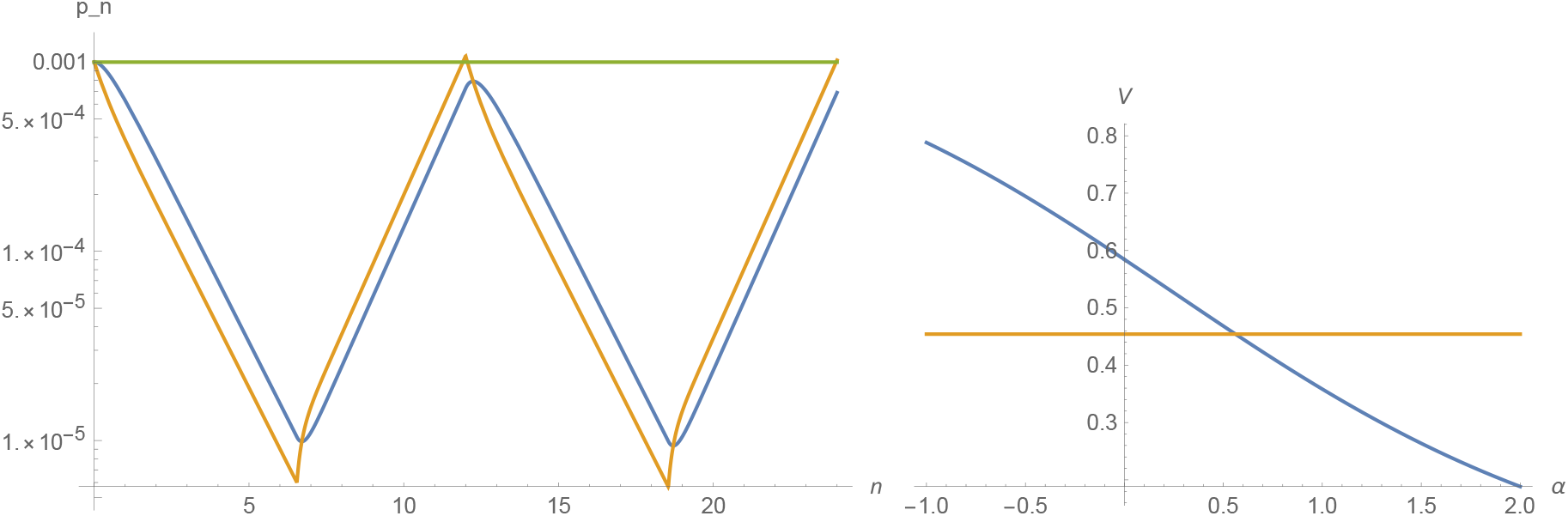
Left: same as Figures 6 and 10 except with Erlang shape parameter 1 instead of 2 or 4, and with cycles of 5.45 weeks up and 6.55 weeks down. Both states shown: *E* (yellow) and *I* (blue). Right: utility normalized as in Figure 5.

## Appendix E Changing incubation and infectious period lengths

The length of the infectious-but-not-isolated period is influenced by policy as well as the underlying dynamics of the disease. In Figure 2 we assumed that mean incubation and infectious-but-not-isolated periods were both 4 days. What would happen if we reduced mean incubation time to 3.5 days and increased mean infectious-but-not-isolated time to 7 days (so that now the mean infectious period is twice the mean incubation period)?

The answer, as illustrated in Figure 12, is that it does not affect the fundamental picture very much if one does this *while holding the R*_EFF_ *values constant*. The tradeoffs evident in Figure 12 are essentially the same as those in Figure 2.

**Figure 12:**
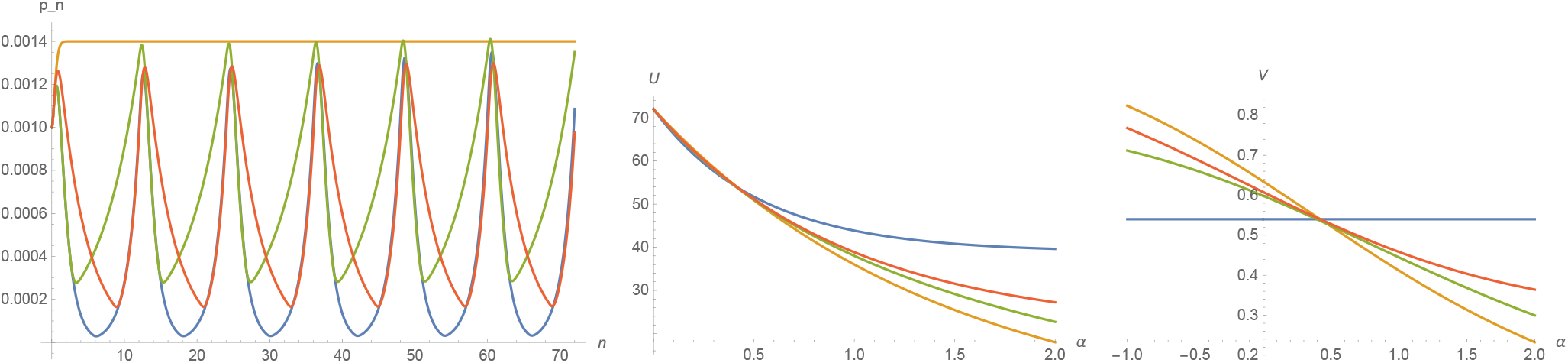
Same as the upper two graphs in Figure 2 (with the alternate utility normalization from Figure 5 on the right) except that mean incubation time is 3.5 and mean infection length is 7. The up-period lengths were reduced slightly to keep the peaks roughly stable: from 6.67 to 6.48 (blue), from 3.6 to 3.5 (green) and from 9.65 to 9.55 (red) and down-period lengths were increased accordingly (so cycle length remains 12 weeks).

But two things are worth pointing out. The first is that Figure 12 begins (like Figure 2) with “equal prevalence in all four states.” But this balance changes quickly even in the steady moderation case, since one now tends to spend twice as much time in the *I* states as the *E* states; so initially more people enter *I* than leave it. That is the reason that all of the curves in the left graph of Figure 12 rise at the beginning. In the *R*_EFF_ = 1 steady state equilibrium, there should be twice as many people in the infectious state as in the exposed state (since it lasts twice as long on average).

The second point to emphasize is that in practice, measures that encourage infected individuals to self-isolate more quickly are fundamentally important *because they decrease R*_EFF_ and changing *R*_EFF_ makes a large difference to required lockdown lengths. It is simply a technical observation that “changing mean infectious-but-not-isolated length while holding *R*_EFF_ constant” seems to have a less pronounced effect.

## Appendix F Lagrange Multipliers and Utility Optimization

We consider different choices for the function *U* and consider what conditions on *U* are necessary in order for intermittent strategies to be preferable to steady moderation.

It will be convenient to work on a logarithmic scale, so we write *b_n_* = - log *c_n_* for the amount that the *negative log* of the infection rate changes (beyond its default change) over the *n*th period. Let *F*(*b*) denote the utility associated to setting *b_n_* = *b* (or equivalently setting *c_n_* = *e*^-^*^b^*). Then

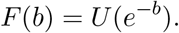

Note that since *U* can be assumed to be an increasing function, *F* will be a decreasing function. For each given *b*, the *marginal cost* of an infinitesimal increase in *b* is given by the negative derivative

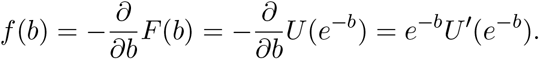

For example, if *U*(*x*) = *x^α^* then *f* (*b*) = *αe*^-^*^b^e*^-(^*^α^*^-1)^*^b^* = *e*^-^*^αb^*. If we imagine *α* = 1 and a hard cutoff at *b* = - ln(1/8) = ln(8) then this could be expressed formally by stating that the marginal cost becomes infinite beyond that point, i.e.,

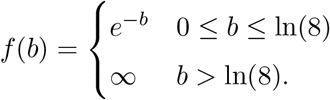

Thus the function *f* starts with *f*(0) = 1 and then decays exponentially until a threshold at which it sharply jumps to ∞. One might argue that this choice of *f* is unrealistic for at least two reasons. First, there are probably certain measures that have low cost relative to their impact. We could account for this “low-hanging fruit” by modifying *f* so that *f* (0) = 0 (while *f* remains otherwise positive and continuous). Second, instead of asserting that it is impossible to go beyond *b* = - ln(1/8) it might be more reasonable to allow *f* to vary continuously but sharply increase beyond that point. Qualitatively, one might see a curve like Figure 13.

**Figure 13:**
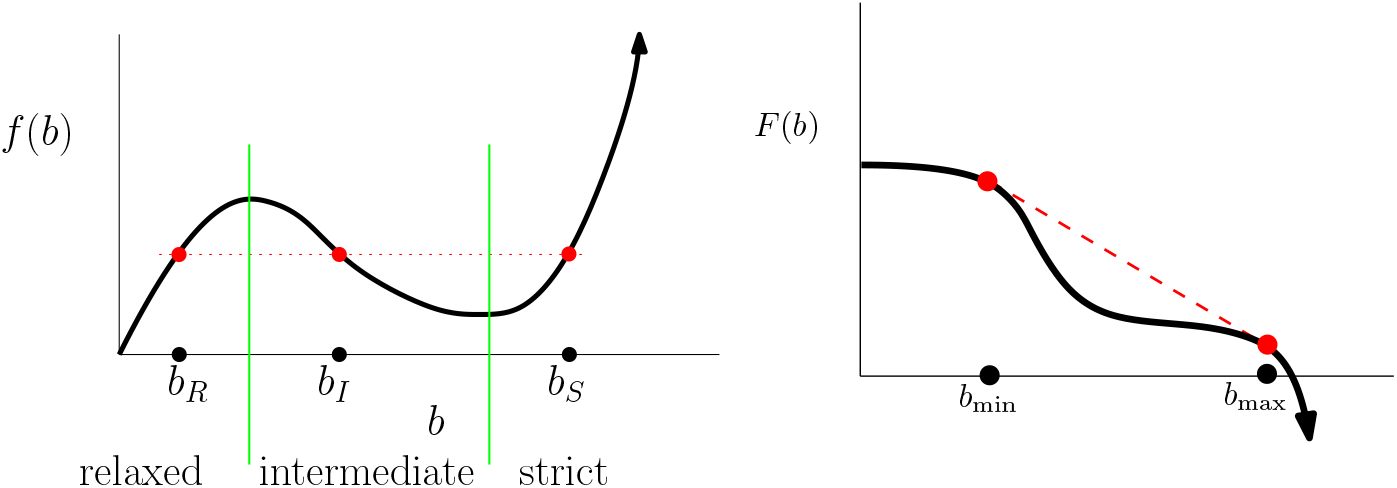
Left: the range of b values is divided into a *relaxed* interval (where *f* is increasing) an *intermediate* interval (where *f* is decreasing) and a *strict* interval (where *f* is increasing again). The values *b_R_*, *b_I_*, *b_S_* shown (one from each region) satisfy *f* (*b_R_*) = *f* (*b_I_*) = *f* (*b_S_*). Right: possible shape for *F*. Recall however that we generally restrict the domain for *F* in order to exclude the very low slope part to the left or the very negatively steep slope part to the right. If we let 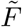 be the smallest concave function satisfying 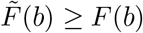 then part of the graph of 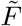 will trace the dotted red line.

However, in Section 2.3 we stated that we would restrict the domain so as to exclude the extreme endpoints (so inexpensive measures like masks hand-washing would stay in place during a *c* = 1 scenario, while unrealistically expensive measures that go beyond *c*_min_ would never be considered). Doing this would amount to recentering so that *b* = 0 corresponded to a location where *f* was positive and that *b*_max_ = - log *c*_min_ would be the largest value we would consider.

Recall also that in Section 2.3 we defined *α*(*c*): = *cU*″(*c*)/*U′*(*c*) and considered three scenarios: convex (*α* > 1), moderate (0 < *α* ≤ 1) and ultra-concave (*α* < 0). Assuming *U* is twice-differentiable, differentiating with the chain rule shows that *U* is ultra-concave at a point c if and only if *f* is concave at *b* = - log *c*, which amounts to *f* having negative first derivative at *b*.

As we have seen in Appendix C, within the linearized SEIR (or SEIS) models, the price of decreasing the number of infections by a certain factor (over the course of a week) actually *depends* on the vector representing the fraction of people in each state at the beginning of the week. To avoid having to account for this in the function *F*, we simply define *F*(*b*) to be the asymptotic long-term cost per time unit of a *steady* policy that decreases the log of the infection number by *b* units during each unit of time (i.e., a policy that has -*b* as the Malthusian parameter, in the language of Appendix C). In other words, *F* does not account for the “switching cost” described in Appendix C. (In Appendix G we will explain how to explicitly derive the *F* corresponding to scenarios like the one in Figure 2, as illustrated see Figure 14.)

**Figure 14:**
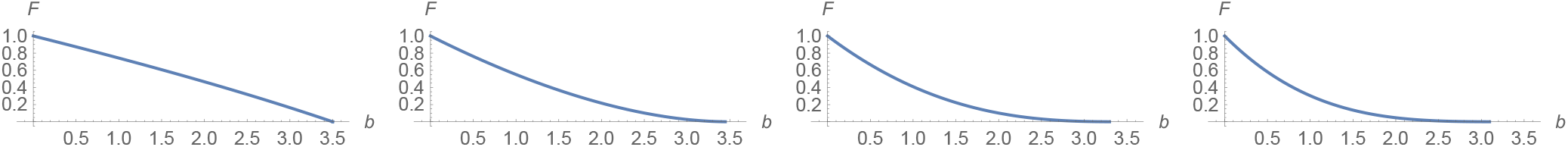
The function *F* corresponding to the disease transmission model from Figure 2 when (from left to right) *α* = .25, *α* = .5, *α* = .75, and *α* = 1. The transition from concave to convex happens somewhere between *α* = .25 and *α* = .5. One can assume that these graphs are horizontally translated so that *b*_min_ corresponds to the point 0.

A natural optimization question can be posed as follows. Suppose policy makers demand that the virus prevalence equal exactly (or at most) some fixed value (say 1/1000) in 12 weeks. Leaving aside the question of total infections for now, how can a constraint like *p*_12_ = 1/1000 be satisfied with the lowest social cost (i.e., the highest *U*)? To address this, note that fixing *p_n_* is equivalent to fixing Σ *b_n_* = *B* for some *B* ≥ 0.

Write *b* = *B/n*. If *b* ϵ [*b*_min_, *b*_max_] then we can consider a strategy that adopts *b*_max_ for a (*b* - *b*_min_)/(*b*_max_ - *b*_min_) fraction of the time *b*_min_ for the remainder of the time. If we define 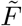 as in Figure 13, then the utility per unit of time would be 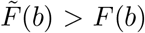 if we ignore the cost of the policy change and we assume that any real length for the time intervals is possible. This is perhaps the clearest way to see why long-term intermittent strategies are beneficial when b lies in an interval on which *F* is convex.

Note that the up-period time duration specified above, namely (*b* - *b*_min_)/(*b*_max_ - *b*_min_), is not necessarily an integer. If we insist that policies be set one week at a time (and that there are *n* weeks total) then we can also use the standard theory of Lagrange multipliers, which says that the minimum of the cost 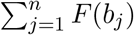 with respect to the constraint 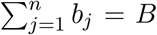 is achieved at a (*b*_1_, *b*_2_,…, *b_n_*) vector for which all *f* (*b_j_*) values are equal to the same value *λ*. (It is not hard to see that unless *B* = 0 this minimum will never be achieved with one of the *b_j_* at the place where *f* is zero, since otherwise shifting that point infinitesimally to right, and any other point infinitesimally left, would reduce the total cost.) Similarly, at most one of the *b_j_* values can lie in the “intermediate range” of Figure 13 where the slope of *f* is negative (since if there were two, one could decrease the cost by adding an infinitesimal amount to one and subtracting it from the other; note that *F* is convex in the relaxed and strict regions but concave in the intermediate region). Thus, for any optimal (*b*_1_,…, *b_n_*) at most one of the *b_j_* can lie in the intermediate region. Although we have been informal, the basic conclusion is this:

### Proposition 1

*Oscillating strategies will appear as optimal solutions to the problem above, for at least some boundary data and n* ≥ 3, *whenever f* (*which we assume to be continuous*) *is decreasing on some interval, or equivalently whenever F is convex on some interval*.

The interesting thing is that whenever *B/n* lies in the intermediate range, the *most costly* choice (among all strategies in which all *b_j_* are in the intermediate range) is the consistent one in which *b_j_* = *B/n* for all *j* ϵ {1, 2,… *n*}.^2^ The *least costly* solution meanwhile involves at most one intermediate *b_j_* — all the other periods are either relaxed or strict. In other words *consistent moderation is* (*among all “intermediate-valued” strategies*) *the worst possible approach* while *the best approach involves solely relaxed and strict periods* (*with at most one intermediate period*). Once we find the optimal Lagrange multiplier solution, we may as well (if we ignore the lower bound on *p_n_*) arrange to put all of the strict periods first and all of the relaxed periods last (so that the trajectory of log *p_n_* follows a V shape, with a possible intermediate period in the middle of the V). The analysis is a bit more complicated if instead of maximizing *U* one aims to maximize *U* - *sP* for some constant *s* > 0 (so that we are taking the infection in account). Although we do not give details here, we note that one might expect that once *P* is taken into account, the log *p_n_* trajectory should lie *below* this V shape, so that there is an even deeper oscillation in this case.

## Appendix G Specific utility function and disease-timing-law examples

In the absence of empirical data on the shape of the utility function, it is important to consider what kinds of functions might be plausible (in “micro” as well as “macro” settings) and what kinds of assumptions underlie the choices. Write *r* = *ψ*(u) for a continuous “disease transmission rate” achieved when the lockdown intensity is chosen so that the utility per week is *u*. Appendix G.1 will present several different possibilities for this function. Our simple model satisfies the simple timing assumption (STA) that an individual who acquires the disease during weak *n* is infectious throughout week *n* + 1 and never infectious thereafter. Under STA, we had *r* = *c* and *U*(*c*) = *ψ*^-1^(c). In Appendix G.2 we will consider what happens if replace STA with other assumptions, such as those that appear in (linearized) SEIR or SEIS models; in these cases the relationship between r and c (the effective growth factor over the course of a whole week) will be a bit more complicated.

In general, *U* should always be an increasing function of c on the interval [0,1] (or at on least some subset corresponding to “plausible” values). We make no claims about how empirically plausible the stories below are, but we hope they help illustrate the ways that different *ψ* functions might be expected in different settings.

### G.1 Other rate functions

1. **Linear policy tool:** *ψ*(*u*) = *u*. Suppose the disease is restricted to children, and our only way to fight the disease is to cancel children’s activities. If *u* is the fraction of activities allowed to take place, we would expect *ψ*(*u*) = *u*.. In this case, we call the cancellation of activities a **linear policy tool** since disease transmission is a linear function of *u*. The closure of factories discussed in Section 2.3 is plausibly (to first order) a linear policy tool.
2. **Quadratic policy tool:** *ψ*(*u*) = *u*^2^. In Section 2.3 we mentioned that if all but a *u* fraction of the population wore perfectly effective masks, then one might expect *ψ*(*u*) = *u*^2^. We hence call the mask policy a **quadratic policy tool**. However, if the masks were only effective in one direction (offering no protection *to* the wearer but perfect protection *from* the wearer) then this would be a linear policy tool. And let us mention one additional subtlety: if the masks were perfectly effective both ways, but it were always the *same* people wearing the masks, then eventually only non-mask-wearers would be infected; at that point the fraction of new infections prevented by masks would be linear in the number of masks.
3. **Intermediate policy tool:** *U*(*c*) = *c^α^*. If a global policy involves a complex mixture of linear and quadratic policies, then one might consider *ψ*(*u*) = *u*^1/^*^α^* for some *α* ϵ [1/2,1] as a compromise.
4. **Convex policy tool:** This story is a bit harder to tell for a constant rate *ψ*(*u*), so let us phrase it terms of *U* and *c*. Suppose a disease only spreads to close friends of infected individuals (and each such individual has at most one infected friend) and that the policy decreases the time *u* that close friends spend together. Then the probability that the friend of an infected individual *remains uninfected* is *e^-ku^* for some *friend transmission rate* k. So we could set *c* = 1 - *e^-ku^* and find *e^-ku^* = 1 - *c* and solve to get and *u* = *U*(*C*) = - log(1 - *c*)/*k*. In this case *U* is convex (meaning that as one adopts measures to decrease *c*, the marginal cost of decreasing c further *decreases*), i.e., this is an example of *accelerating returns*. This function only makes sense for *c* ϵ [*a*, 1] for some *a* < 1, since there is an upper bound to the time friends can spend together. A similar phenomenon appears in the discussion of SEIR on a three-regular tree in Section G.2.

**Remark on** *α* = 1 **versus** *α* = 1/2: In the context of SEIR, the infection rate is said to be proportional to *SI/N*. If we formally interpret a “lockdown” as a measure that temporarily decreases *S* and *I* (by a factor proportional to utility) then the lockdown would be a quadratic tool, as in [23]. If we formally interpret a lockdown as a measure that temporarily decreases *S*, *I*, and *N* by the same factor then it would be a linear tool. It is not clear what the correct answer *should* be because it depends on whether non-locked-down people continue to have the usual number of contacts per day or have fewer because there are fewer people for them to interact with. Sample empirical question: if one closes half the bars in a town and locks down half the customers, what do the other customers do? Do half of the non-locked-down customers stay home—because their favorite bar is closed—or do all non-locked-down customers find open bars and have as much social contact as usual? In the former case, how is the absence of the non-locked-down customers who stay home accounted for in the utility function? Are they “effectively locked down”? If it is the *same* customers locked down each week, then after a couple of weeks, most of the increase in cases would come from the non-locked-down population and (if they were interacting with each other at normal rates) the overall growth rate might be similar to what it would be if there were no measures in place at all. To model an “inhomogeneous” scenario like this, one could to introduce separate compartments (e.g., for essential and non-essential workers) but we will not do this here.

### G.2 The timing of the infectious period and power law utility

We will now explicitly derive *U* and *F* in a few very simple examples. Recall that when translated into probabilistic language, the SIR model effectively assumes that there is no incubation period and that the disease duration is an exponential random variable. In SEIR (with Erlang parameter 1) the incubation period is also an exponential random variable. As a simple example in this section we will consider a linearized version of SEIR/SEIS in which the incubation time has Erlang parameter 2 (so *E* is divided into two states *E*_1_ and *E*_2_) but the infectious time is exponential (so *I* is not divided into two states).

One (possibly unrealistic) aspect of all three of these models is that there is *no lower bound* on the incubation period, which in principle means there is no upper limit to the number of “hops” a virus can make during a unit of time. This allows for a type of “continuous compounding” that does not appear when the incubation time is bounded below. SIR has incubation times of length zero, but short incubation periods are less likely in SEIR when the exposed state has Erlang parameter 1, and even less likely when the Erlang parameter is higher. On the other hand, STA assumes (perhaps unrealistically) that there is an *upper bound* on the length of time an individual is infectious but not quarantined (although one could alternatively reinterpret the lower bound on c as corresponding to a positive fraction of infectious individuals who remain infectious for the next week).

We now consider these four timing rules (STA, SIR, SEIR, Erlang SEIR) along with an additional example involving SEIR on a three-regular tree; and by way of illustration we will check explicitly how they effect the calculation of *U* in the power law case where *r* = *ψ*(*u*) = *u*^1/^*^α^* and *α* > 0.

In the examples below, we will interpret *c* as the exponential of the *Malthusian parameter* of the given disease dynamics. Since the Malthusian parameter describes the asymptotic *continuous* exponential growth rate, its exponential is the growth factor over a single period. In contrast to the first model in the paper, the examples below will take *c* (rather than 2.5*c*) to represent the (asymptotic) weekly growth factor *p_n_*_+1_/*p_n_* corresponding to a given strategy. This simplifies the formulas slightly, and also means that in order to calculate *U*(*c*) we will not necessarily need to specify the minimal and maximal growth factors *c*_min_ and *c*_max_.

1. **Simple Timing Assumptions (STA):** The conditions presented at the beginning of this paper were obviously contrived to allow for a simple discretization of the problem. On the other hand, these conditions have two features worth highlighting: first a lag time (of some part of a week) between exposure and infectiousness. And second an upper bound on the length of time an individual is able to be infectious. Recall that with this duration law we have:

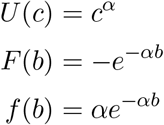 Note here that *c^α^* and (2.5*c*)*^α^* agree up to a constant factor, so we can still use *c^α^* as the expression for *U*(*c*) now that we are using *c_n_* instead of 2.5 *c_n_* to represent *p_n_*_+1_/ *p_n_*. Note also that *f* is a strictly decreasing function for *any α* > 0. Recalling Proposition 1, this means that under STA and any power law *ψ*, it is *always* the case that optimal strategies alternate between the most extreme allowable values.
2. **Linearized SIR or SIS:** The classical SIR model can be derived from the assumption that an individual who catches a virus is instantly infectious, with an infection duration that is an exponential random variable. If we take the *S* → ∞ limit, we obtain a linearized version of the model in which no herd immunity develops and *I* grows exponentially, with a rate given by a difference between the infection rate and the recovery rate. If *I* starts out at 1 at time 0, then its value at time 1 is the exponential of an affine function of *r*. To choose an arbitrary example, say *c* = *e^r^*^-1^. Then *r* = 1 + log(*c*), and since *u* = *r^α^* we obtain

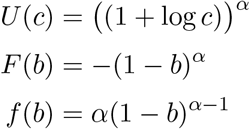

on the range of plausible *c* values. In this case *f* is increasing only if *α* > 1.
3. **Linearized SEIS or SEIR:** The standard SEIR model can be derived from the assumption that the exposed and infectious periods for an individual are independent exponential random variables. Once again we can take the *S* → ∞ limit to obtain a linearized version of the model in which no herd immunity develops, and we can discretize it by assuming that policy is set one week at a time. In this case *E* and *I*, interpreted as functions of *t*, evolve according to a linear ODE. Even this linearized version of SEIR is more complicated than the linearized version of SIR above because it has two parameters to keep track of instead of 1. If we let *β* denote the infection rate (and set other parameters equal to one, for illustration) we obtain

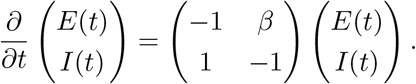 The standard theory of linear ODEs says that if we denote the above matrix by *M_β_*, and we start with *E* = *E*_0_ and *I* = *I*_0_, then the values at time *t* will be given by

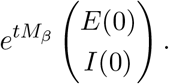 The value of *E*(*t*) + *I*(*t*) at time *t* depends on both *E*_0_ and *I*_0_. But regardless of the initial values, as t gets larger, standard linear algebra implies that this quantity grows asymptotically like a constant times *e^λt^* where *λ* = *λ_β_* is the largest eigenvalue of *M_β_*. This *λ_β_* is called the *Malthusian parameter* as we mentioned earlier, see [11]. We can quickly compute the eigenvalues (type Eigenvalues({{-1,b},{1,-1}}) into wolframalpha.com) and the matrix *e^M^* (type matrixexp(t {{-1,b},{1,-1}}) into wolframalpha.com) and we find that the largest eigenvalue of *M* is 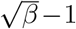. Write A(*t*) = E(*t*) +1(*t*) for the total virus carriers at stage t if the parameter *β* remains fixed. A glance at the matrix shows that if *A*(*t*) = *E*(*t*) + *I*(*t*) then regardless of what *E*(*t*) and *I*(*t*) are, the value *A*(*t*) will be within some fixed constant factor of 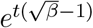 for all *t*. In order to avoid having to think too hard about non-commutative matrices (and the “switching costs” discussed in Figure 3 and Section C), let us assume that the constant-*β* periods we consider are long (as in the V shape considered Section F). If this is the case, then the *effective* multiplicative factor at each step is (very close to) 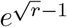 where *r* = *β*. Taking this point of view puts us back in a similar framework to the (linearized) SIR problem. Solving we find *r* = (1 + log *c*)^2^ and combining this fact with *u* = *r^α^* we obtain the following (defined on the interval where (1 + log *c*) is positive):

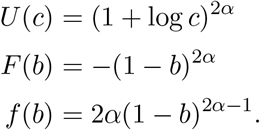 Although we have not worked out the *very optimal* solution, the above is enough to show that if *f* is decreasing over its range (as it is when *α* > 1/2) then there is a “strict-then-relaxed” solution (analogous to the V-shaped one in Section F) that is better than a solution in which *β* is held constant throughout. In other words alternating strict then relaxed beats steady moderation (at least over sufficiently long periods—long enough so that the “switching costs” detailed in Section C are small compared to the total costs).
4. **Linearized SEIS or SEIR** with Erlang parameter 2 for exposed state: To illustrate one more example, consider a type of Erlang SEIR in which each individual who acquires the virus passes through two exposed states before reaching a single infectious state. In such a setting, the total length of the incubation period is the *sum* of two independent exponential random variables. For example, if *X*_1_ and *X*_2_ are independent exponential random variables, each with density function *e*^-^*^t^* on [0, ∞), then their sum has density function *te*^-^*^t^* on [0, ∞). The sum is called an Erlang (or gamma) random variable, and the fact that the density is zero at zero encodes the fact that “extremely short” incubation periods are unlikely. (A similar gamma random variable is used to model the incubation period of the flu in [37].) Erlang SEIR is still a simplified example, but as noted earlier its incubation period law may be more realistic than the one with Erlang parameter 1. As before, when we take the *S* → ∞ limit we obtain a linearized version of the model which follows a linear ODE where *β* denotes the infection rate (and we set other parameters equal to one, for illustration):

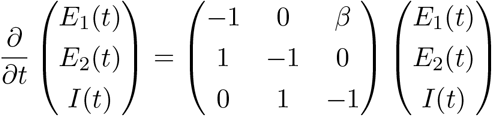

and if we type Eigenvalues[{{-1, 0, b}, {1, −1, 0}, {0, 1, −1}}] into wolframalpha.com we find the largest eigenvalue is 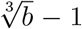. So setting *β* = *r* and following the same analysis as in the previous example we get 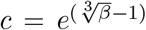. Solving we find *r* = (1 + log *c*)^3^ and combining this with *u* = *r^α^* we obtain the following (defined on the interval where (1 + log *c*) is positive):

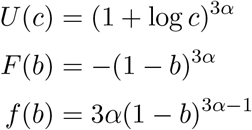 In this case *f* is increasing provided *α* > 1/3. Although we have only worked out only simplified cases in this section, the idea that *α* = 1/3 is the cutoff for linearized SEIR/SEIS (with Erlang parameter 2) and *α* = 1/2 for ordinary SEIR (with Erlang parameter 1) seems *roughly* in line with Figures 2 and 11. It is also not surprising that higher (and possibly more realistic) Erlang parameters lead to intermittent strategies being optimal for even smaller values of *α*, as in Figure 10.
5. **Linearized SEIR on a three-regular tree:** When a disease spreads mainly among close associates of infected individuals, it can in principle lead to *U* being convex (or at least somewhat less concave). We include here an analytically simple example (not intended to be realistic) to illustrate that point. Imagine a scenario in which each individual has exactly three very close associates (e.g., a work colleague, a spouse, and one other friend). For simplicity, let us imagine that the associate graph does not have short cycles, so that it looks locally like a three-regular tree. (Allowing some short cycles would not necessarily change the basic story, but it would make the math more complicated.) As the disease spreads on the tree, we will keep track only of the number of infected individuals who have at least one susceptible neighbor (since these are the individuals who could still spread the disease to others). Precisely, we consider three states: exposed with two susceptible neighbors (*E*_2_), infected with two susceptible neighbors (*I*_2_), and infected with one susceptible neighbor (*I*_1_), evolving according to the ODE

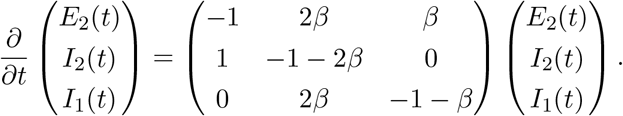 A key thing to note here is that (unlike in the linearized SEIR examples above) no matter how large *β* is, there is an upper bound to how fast the disease can spread: if *β* is very close to infinity, then of the people in the three categories above, nearly everyone will be in the *E*_2_ state (since as soon one transitions from *E*_2_ to *I*_2_, one almost immediately exposes two more neighbors and eliminates oneself, effectively increasing *E*_2_ by 1) and *E*_2_ grows with exponential rate close to 1. Setting *β* = *r*, we enter `Max[Eigenvalues{{-1,2r, r},{1,-1-2r,0},{0,2r, -1-r}}]` into wolframalpha.com and find the largest eigenvalue is 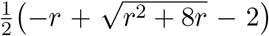. As above, we set 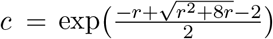. Entering `Solve[c = Ê(.5 (-r + sqrt(r̂2 + 8r) - 2)), r]` we then find

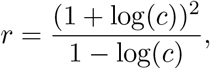

and combining this with *u* = *r^α^* we obtain the following (defined on the interval where *e*^-1^ < *c* < *e*, or equivalently where -1 < *b* < 1):

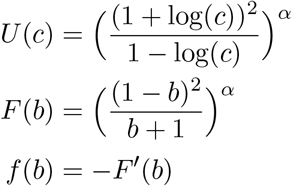 The function *U* is then convex for a range of *α* values, as Figure 15 shows. (Note that we have not specified values for *c*_min_ and *c*_max_.)

**Figure 15:**
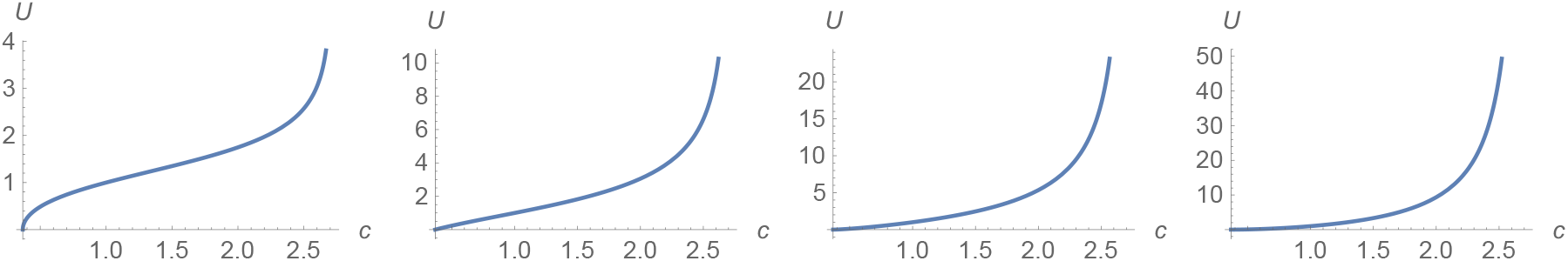
*U*(*c*) for 3-regular-tree SEIR when (from left to right) *α* = .25, *α* = .5, *α* = .75, and *α* = 1.

Finally, we remark that the examples above can be generalized to involve any number of *E* and *I* states, and any discrete or continuous time Markov chain [11], as well as other types of differentiated compartments (e.g., accounting for different disease phases or different demographic categories).

### G.3 Linearized SIR vs. STA in a simple example

To illustrate the distinction between timing laws further, suppose (in a somewhat extreme scenario) that there are 400 infectious people, and that with no intervention the number would quadruple to 1600 in a week, but that with the maximum possible intervention it would decline to 100. Define a “half measure” to be an intervention half as costly as the maximum. How many infectious people would there be after a half measure?

Under STA with a linear policy tool, the answer is 850 (the arithmetic mean of 100 and 1600). Under (linearized) SIR with a linear policy tool, the answer is 400 (the geometric mean of 100 and 1600), because in this case it is the “exponential growth rate” that is linear in cost. Under (linearized) SIR with a quadratic policy tool the answer is 200. Under STA with a convex policy tool, the answer would be some value *greater* than 850.

## Appendix H Mathematica code

The reader who wants to experiment with different parameters may cut and paste the following code into Mathematica, which generates Figure 6. Other figures in this paper use similar code. This code is not optimized for computational efficiency (since Mathematica presumably computes the matrix exponentials separately for each point it plots) and researchers using much larger matrices (to represent models with more compartments) may wish to solve the ODEs a different way. To parse the code, note that the first two lines instruct Mathematica to always interpret the 0th power of a matrix as the identity, which is not true by default in Mathematica when a matrix is (nearly) singular. The Jacobian matrix is then written as *m* (a function of the effective *R*_0_ parameter, here called *R*) and the exponential as *M* = *e^tm^* (a function of R and t). These matrices are applied on the left to column vectors, and the initial vector is written as {1,1,1,1} (multiplied by .001). Note that typing *N*[*X*] instructs Mathematica to produce a numerical approximation of *X*, that Re[*X*] is the real part of *X* (which eliminates some small complex-valued numerical errors that arise in the matrix operations) and that the period symbol is used to denote both matrix multiplication and dot product.

`Unprotect[MatrixPower]; MatrixPower[m_?SquareMatrixQ, 0]:=IdentityMatrix[Length[m]];`

`Protect[MatrixPower];`

periods = 2; weeksup = 6; weeksdown = 6; Rup = 2.2; *R*_DOWN_ = .33; incubationdays = 4; infectiousdays = 4;

`m[R_]: = {{-14/incubationdays, 0, 7 R/infectiousdays, 7 R/infectiousdays},`

`{14/incubationdays, -14/incubationdays, 0, 0},`

`{0, 14/ incubationdays, -14/infectiousdays, 0},`

`{0, 0, 14/infectiousdays, -14/ infectiousdays}};`

`M[R_, t_] = Re[N[MatrixExp[t m[R]]]]; len = weeksup + weeksdown;`

MFullPeriod = M[Rup, weeksup].M[*R*_DOWN_, weeksdown];

MPartPeriod[r_] = If[r < weeksdown/len, M[*R*_DOWN_, r len], M[Rup, r len - weeksdown].M[*R*_DOWN_, weeksdown]]; LogPlot[{

`.001 N[{0, 0, 0, 1}.MPartPeriod[a/len - Floor[a/len]].MatrixPower[MFullPeriod, Floor[a/len]].{1, 1, 1, 1}],`

`.001 N[{0, 0, 1, 0}.MPartPeriod[a/len - Floor[a/len]].MatrixPower[MFullPeriod, Floor[a/len]].{1, 1, 1, 1}],`

`.001 N[{0, 1, 0, 0}.MPartPeriod[a/len - Floor[a/len]].MatrixPower[MFullPeriod, Floor[a/len]].{1, 1, 1, 1}],`

`.001 N[{1, 0, 0, 0}.MPartPeriod[a/len - Floor[a/len]].MatrixPower[MFullPeriod, Floor[a/len]].{1, 1, 1, 1}]}, {a, 0, len*periods}, Exclusions -> None, AxesLabel -> {n, p_n}, ImageSize -> 600]`

## Footnotes

1 The arithmetic-geometric mean inequality implies that for any choice of *α* > 0 “steady moderation” (i.e., maintaining *R*_EFF_ = 1 over a long period) results in a lower *U* value than *any* variable program that achieves the same final *p_n_* value. More generally, if *U* is either convex or moderate on some interval (*a,b*), then keeping *c_n_* equal to a constant in this interval over an extended period is more costly (in both utility and infection rates) than alternating between *c_n_* ≤ *α* and *c_n_* ≥ *b*. Appendix F explains more generally the mathematical properties of *U* that cause this behavior.

2 This fact is known in mathematics as *Jensen’s inequality*.

## References

[1] Ensheng Dong, Hongru Du, and Lauren Gardner. An interactive web-based dashboard to track COVID-19 in real time. The Lancet Infectious Diseases, 2020.

[2] Latinos in Some States Have Seen Higher Rates of Infection. The New York Times, May 2020.

[3] Neil M Ferguson, Daniel Laydon, Gemma Nedjati-Gilani, Natsuko Imai, Kylie Ainslie, Marc Baguelin, Sangeeta Bhatia, Adhiratha Boonyasiri, Zulma Cucunubá, Gina Cuomo-Dannenburg, et al. Impact of non-pharmaceutical interventions (NPIs) to reduce COVID-19 mortality and healthcare demand. Imperial College, London. DOI: https://doi.org/10.25561/77482, 2020.

[4] Stephen M Kissler, Christine Tedijanto, Marc Lipsitch, and Yonatan Grad. Social distancing strategies for curbing the COVID-19 epidemic. *medRxiv*, 2020.

[5] Dylan H Morris, Fernando W Rossine, Joshua B Plotkin, and Simon A Levin. Optimal, near-optimal, and robust epidemic control. *arXiv preprint arXiv:2004.02209*, 2020.

[6] Fernando Alvarez, David Argente, and Francesco Lippi. A simple planning problem for covid-19 lockdown. *medRxiv*, 2020.

[7] Daron Acemoglu, Victor Chernozhukov, Iván Werning, and Michael D Whinston. A Multi-Risk SIR Model with Optimally Targeted Lockdown. Working Paper 27102, National Bureau of Economic Research, May 2020. Series: Working Paper Series.

[8] Omer Karin, Yinon M Bar-On, Tomer Milo, Itay Katzir, Avi Mayo, Yael Korem, Boaz Dudovich, Eran Yashiv, Amos J Zehavi, Nadav Davidovich, et al. Adaptive cyclic exit strategies from lockdown to suppress covid-19 and allow economic activity. *medRxiv*, 2020.

[9] Michelangelo Bin, Peter Cheung, Emanuele Crisostomi, Pietro Ferraro, Connor Myant, Thomas Parisini, and Robert Shorten. On fast multi-shot epidemic interventions for post lock-down mitigation: Implications for simple covid-19 models. *arXiv preprint arXiv:2003.09930*, 2020.

[10] Helen Abbey. An examination of the Reed-Frost theory of epidemics. Human biology, 24(3):201, 1952.

[11] Odo Diekmann, JAP Heesterbeek, and Michael G Roberts. The construction of next-generation matrices for compartmental epidemic models. Journal of the Royal Society Interface, 7(47):873–885, 2010.

[12] Sheng Zhang, MengYuan Diao, Wenbo Yu, Lei Pei, Zhaofen Lin, and Dechang Chen. Estimation of the reproductive number of novel coronavirus (covid-19) and the probable outbreak size on the diamond princess cruise ship: A data-driven analysis. International Journal of Infectious Diseases, 93:201–204, 2020.

[13] Shi Zhao, Qianyin Lin, Jinjun Ran, Salihu S Musa, Guangpu Yang, Weiming Wang, Yijun Lou, Daozhou Gao, Lin Yang, Daihai He, et al. Preliminary estimation of the basic reproduction number of novel coronavirus (2019-ncov) in china, from 2019 to 2020: A data-driven analysis in the early phase of the outbreak. International journal of infectious diseases, 92:214–217, 2020.

[14] Ying Liu, Albert A Gayle, Annelies Wilder-Smith, and Joacim Rocklöv. The reproductive number of covid-19 is higher compared to sars coronavirus. Journal of travel medicine, 2020.

[15] Xi He, Eric HY Lau, Peng Wu, Xilong Deng, Jian Wang, Xinxin Hao, Yiu Chung Lau, Jessica Y Wong, Yujuan Guan, Xinghua Tan, et al. Temporal dynamics in viral shedding and transmissibility of covid-19. Nature medicine, pages 1-4, 2020.

[16] Chaolong Wang, Li Liu, Xingjie Hao, Huan Guo, Qi Wang, Jiao Huang, Na He, Hongjie Yu, Xihong Lin, An Pan, et al. Evolving epidemiology and impact of non-pharmaceutical interventions on the outbreak of coronavirus disease 2019 in Wuhan, China. *medRxiv*, 2020.

[17] Chloe Taylor. How New Zealand’s ‘eliminate’ strategy brought new coronavirus cases down to zero, May 2020. Library Catalog: www.cnbc.com Section: Health & Science.

[18] Adam J Kucharski, Timothy W Russell, Charlie Diamond, Yang Liu, John Edmunds, Sebastian Funk, Rosalind M Eggo, Fiona Sun, Mark Jit, James D Munday, et al. Early dynamics of transmission and control of covid-19: a mathematical modelling study. The lancet infectious diseases, 2020.

[19] https://www.youtube.com/watch?v=V0lfVur2UWA&feature=emb_logo. 2020 (accessed April 22, 2020).

[20] Joseph A Lewnard, Vincent X Liu, Michael L Jackson, Mark A Schmidt, Britta L Jewell, Jean P Flores, Chris Jentz, Graham R Northrup, Ayesha Mahmud, Arthur L Reingold, et al. Incidence, clinical outcomes, and transmission dynamics of hospitalized 2019 coronavirus disease among 9,596,321 individuals residing in california and washington, united states: a prospective cohort study. *medRxiv*, 2020.

[21] Maria Do Rosário De Pinho, Igor Kornienko, and Helmut Maurer. Optimal control of a SEIR model with mixed constraints and l 1 cost. In CONTROLO’2014-Proceedings of the 11th Portuguese Conference on Automatic Control, pages 135-145. Springer, 2015.

[22] Md Haider Ali Biswas, Luís Tiago Paiva, and MDR De Pinho. A SEIR model for control of infectious diseases with constraints. Mathematical Biosciences and Engineering, 11(4):761–784, 2014.

[23] Fernando Alvarez, David Argenta, and Francesco Lippi. A simple planning problem for COVID-19 lockdown. https://sites.google.com/site/dargenteamaya/research, 2020 (accessed March 29, 2020).

[24] Fast Facts on U.S. Hospitals, 2020 | AHA. Library Catalog: www.aha.org.

[25] Jomar F. Rabajante. Insights from early mathematical models of 2019-nCoV acute respiratory disease (COVID-19) dynamics, 2020.

[26] Zhihua Zhu, Jiansen Li, Dexin Gong, Donghua Wan, Shaowei Chen, Lingchuan Guo, Yan Li, Limei Sun, Wenjia Liang, Tie Song, et al. Time-varying transmission dynamics of novel coronavirus pneumonia in China [j]. 2020.

[27] Julien Arino and Stéphanie Portet. A simple model for COVID-19. Infectious Disease Modelling, 2020.

[28] Stephen A Lauer, Kyra H Grantz, Qifang Bi, Forrest K Jones, Qulu Zheng, Hannah R Meredith, Andrew S Azman, Nicholas G Reich, and Justin Lessler. The incubation period of coronavirus disease 2019 (COVID-19) from publicly reported confirmed cases: estimation and application. Ann Intern Med, 10:M20–0504, 2020.

[29] Liangrong Peng, Wuyue Yang, Dongyan Zhang, Changjing Zhuge, and Liu Hong. Epidemic analysis of COVID-19 in china by dynamical modeling. *arXiv preprint arXiv:2002.06563*, 2020.

[30] Richard A Neher, Robert Dyrdak, Valentin Druelle, Emma B Hodcroft, and Jan Albert. Potential impact of seasonal forcing on a sars-cov-2 pandemic. Swiss Medical Weekly, 150(1112), 2020.

[31] Mohammad M Sajadi, Parham Habibzadeh, Augustin Vintzileos, Shervin Shokouhi, Fernando Miralles-Wilhelm, and Anthony Amoroso. Temperature and latitude analysis to predict potential spread and seasonality for covid-19. *Available at SSRN 3550308*, 2020.

[32] Jingyuan Wang, Ke Tang, Kai Feng, and Weifeng Lv. High temperature and high humidity reduce the transmission of covid-19. *Available at SSRN 3551767*, 2020.

[33] Alexandra A. Sidorenko and Warwick J. https://www.brookings.edu/opinions/what-a-flu-pandemic-could-cost-the-world/ McKibbin. what a Flu Pandemic Could Cost the World, 2009 (accessed April 3, 2020).

[34] Warwick J. McKibbin and Roshen Fernando. The global macroeconomic impacts of COVID-19: Seven scenarios. 2020 (accessed April 3, 2020).

[35] Wen Hai, Zhong Zhao, Jian Wang, and Zhen-Gang Hou. The short-term impact of SARS on the chinese economy. Asian Economic Papers, 3(1):57–61, 2004.

[36] Anu G Gupta, Cheryl A Moyer, and David T Stern. The economic impact of quarantine: SARS in Toronto as a case study. Journal of Infection, 50(5):386–393, 2005.

[37] Hiroshi Nishiura, Nick Wilson, and Michael Baker. Quarantine for pandemic influenza control at the borders of small island nations. BMC infectious diseases, 9:27, 04 2009.

